# Chest Wall Restriction Device for Modeling Respiratory Challenges and Dysfunction

**DOI:** 10.1101/2025.01.08.24319742

**Authors:** Victoria Ribeiro Rodrigues, Lizuannette Mejia, Rafael G. Zucchi, Paul W. Davenport, Nicholas J. Napoli

## Abstract

Breathing relies on unrestricted movement of the chest wall to maintain O2 and CO2 balance. Understanding the effects of chest and abdominal restrictions on respiratory function is critical for studying conditions such as respiratory diseases, extreme environments, and load-induced impairments. However, existing methods to simulate these restrictions are limited, lacking the ability to provide both static and dynamic conditions or precise load control. To address these gaps, we developed a novel chest wall and abdomen restriction device capable of independently applying and measuring static and dynamic loads with adjustable and reproducible force levels. Separate bands for the chest and abdomen enable targeted restrictions, with constant force springs providing resistance during dynamic conditions and immobilization during static conditions. Integrated sensors quantify applied loads and respiratory mechanics. To validate the device, healthy participants underwent pulmonary function testing under baseline, static, and dynamic restriction conditions. Significant reductions in forced expiratory volume (FEV1) and forced vital capacity (FVC) were observed under restrictions compared to baseline. Other respiratory metrics also differed significantly, highlighting distinct effects of static and dynamic restrictions. Pressure variability tests confirmed reproducibility and adjustability of loads, while displacement data from linear variable differential transducers (LVDTs) validated the device’s ability to distinguish static and dynamic effects. This device addresses prior limitations by enabling precise, reproducible loading and independent control of chest and abdominal restrictions, supporting research into respiratory diseases, extreme environments, and respiratory mechanics. Our results demonstrate its potential to advance respiratory function research and expand clinical and experimental applications.

## I. Introduction

Breathing is an essential activity performed by the human body, serving as the cornerstone for all other activities. When one is unable to breathe, it disrupts the balance between oxygen (O_2_) and carbon dioxide (CO_2_) in the blood [1]. Therefore, the human body initiates a cascade of effects, leading to a lack of attention, confusion, loss of consciousness, morbidity, and mortality [2], [3], [4]. The chest wall plays a crucial role in the breathing process, serving as the housing for the lungs and dynamically expanding and contracting to facilitate air exchange [5]. Thus, any restriction in the chest wall disrupts the breathing mechanism [6], [7], [5]. These restrictions may originate from physiological factors affecting thoracic compliance and muscle function, such as scoliosis, muscular dystrophy, and diaphragmatic paralysis [8], [9], [10], [11], or from environmental factors, such as high g-forces, water pressure during deep-sea diving, compression from tight clothing like corsets or body armor, or heavy backpacks and safety harnesses used in activities like hiking, rock climbing, or skydiving [12], [13], [14], [15]. However, studying these conditions is challenging due to the limited availability of patients, the logistical complexity of recreating environmental factors, and the need for controlled, repeatable experiments to ensure reliable data. To overcome these issues, we have developed a novel device designed to mimic chest and abdominal restrictions, enabling the exploration of both static and dynamic load-based restrictions. This novel device provides a controlled environment to study the effects of chest and abdominal restriction on breathing.

### A. Prior Work

We can categorize chest wall restrictions into two distinct types: 1) static restrictions and 2) dynamic load-based restrictions. In the static condition, the chest wall is immobile regardless of the force applied [16]. Previous attempts to restrict the chest wall have mostly focused on static restriction. One of the earliest designs was a simple chest clamp used to limit the movement of the rib cage. The device consisted of two plywood boards—one placed on the posterior surface and the other on the anterior surface of the thorax [17]. This approach, however, fails to match the contour of the body, restricting the rib cage only along the sternum and the spine. A more effective alternative that addresses this limitation is chest wall strapping (CWS). This procedure involves restricting the thorax and abdomen using fabric, cinch straps, and/or a chest cast [18], [19], [20], [21], [22]. While CWS is used today, it lacks the precision to quantify the load being added to the chest wall. Furthermore, none of these methods have a metric to allow for reproducible load levels or differentiate between chest and abdominal restriction.

In the dynamic condition, a consistent load is applied to the chest wall, allowing movement only when the participant generates sufficient force to overcome the resistance [16], [23], [24], [25]. Previous efforts to design a dynamic restriction device that maintains the load while permitting moderate movement include a hard-shell, full-body apparatus. This device altered the pressure exerted on the chest in proportion to the inspired volume [26]. However, it required participants to remain in a supine position and lacked a mechanism to quantify the applied load. Another attempt involved a fiberglass chest cast equipped with inflatable air pockets that allowed for controlled load adjustments [16]. Despite this innovation, the device failed to restrict the abdomen and did not provide a way to measure the participant chest and abdomen movement. Finally, none of these devices offer the capability to provide both static and dynamic restrictions within a single design.

These gaps left by the previous designs lead to the following research questions:

RQ1 Can a single device be engineered to integrate both static and dynamic restriction capabilities, enabling comprehensive investigation of their distinct effects on respiratory mechanics?

RQ2 Can such a device be designed to deliver reproducible and precisely adjustable loads to both the chest and abdomen, facilitating targeted and reliable experimentation?

### B. Challenges

Designing a chest wall restriction device capable of simulating a wide range of restrictions presents significant challenges. While existing methods in the literature often focus on replicating specific conditions, such as simulating respiratory diseases like cystic fibrosis or external loads from equipment like bulletproof vests, they are typically limited in scope. Previous methods lacked the precise control needed to adjust the load or the ability to transition between static and dynamic restrictions, making it difficult to tackle the diverse range of physiological and environmental conditions. Developing a more versatile device requires not only precise control over these variables but also the ability to accurately replicate both static and dynamic restrictions to better mimic real-world scenarios. Furthermore, prior methods lacked the ability to accurately quantify the applied load, as they did not have sensors or mechanisms to measure the precise force exerted on the chest wall. Consequently, researchers could not replicate specific conditions or measure the impact of varying load levels with accuracy.

Additionally, these methods did not differentiate between restrictions applied to the chest and those applied to the abdomen. These tools were often built as single-piece or full-coverage restraints, designed for general chest compression rather than targeted restriction, limiting the ability to isolate effects between the chest and abdomen. Finally, the inability to differentiate between chest and abdominal restrictions, combined with the use of single-piece designs, made measurement of chest and abdominal movement during restriction unfeasible. This limitation prevented detailed analysis of how restricted movement influences respiratory dynamics, making it difficult to capture or quantify subtle changes in breathing patterns.

### C. Insights

To address these challenges, we developed a novel restriction device that uses two separate bands, one for the chest wall and one for the abdomen, allowing us to restrict each area independently. In the static condition, both bands are firmly held in place, preventing any movement. This setup simulates scenarios where the chest and abdomen are entirely restricted, similar to real-life situations requiring external braces or devices used to immobilize the torso. For the dynamic condition, the bands that restrict the chest and abdomen are held in place by a constant force spring, allowing the participant to move their chest and abdomen if they apply enough force to overcome the spring resistance. This design enables us to study the effects of dynamic loads where movement is possible but resisted, simulating conditions like respiratory diseases (e.g., COPD) or high G-force environments encountered by jet pilots or race car drivers [27]. Additionally, two Linear Variable Differential Transducers (LVDTs) measure the horizontal displacement of the chest wall and abdomen during breathing under dynamic load, enabling us to detect and quantify subtle changes in breathing patterns.

The device’s vinyl-like fabric bands conform to the body, ensuring even load distribution across each targeted area. The inclusion of constant force springs enables the precise application of a known load to the chest and abdomen. Two pressure sensors monitor the pressure exerted by the bands, ensuring that the applied load is not only accurately set but also consistently maintained across different participants and trials. A calibration process ensures the device provides reproducible and adjustable loading conditions for a variety of experimental setups. By combining controlled loading, precise displacement measurement, and pressure monitoring, our device not only enables the study of static and dynamic chest wall restrictions on breathing but also provides a versatile platform to investigate their effects on cognitive function, blood circulation, and other physiological responses in a controlled environment [28], [29].

### D. Contributions

Our chest wall and abdomen restriction device is a powerful tool for studying respiratory challenges and dysfunction. Combining static and dynamic restrictions in one design allows us to simulate full movement restriction or use adjustable dynamics loads, creating a controlled environment to study how breathing adapts under different conditions. With separate bands for the chest and abdomen, we can isolate how each region contributes to breathing patterns, and built-in sensors like LVDTs allow us to monitor even the smallest changes in movement. The device is also highly reliable, using constant force springs and pressure sensors to ensure precise and repeatable restrictions. This makes it ideal for exploring respiratory diseases, the cognitive effects of breathing restrictions, and how the body responds to extreme conditions, opening up new possibilities for research and treatment.

## II. Methods and Materials

### A. Structure Design

This section provides a detailed description of the different parts of the device. For a comprehensive list of all components used, including detailed dimensions, refer to the schematics in Appendices A-R. All parts were sourced from McMaster-Carr, except for the LVDTs, which were supplied by Omega.

#### 1) Structure and Frame

The chest wall and abdomen restriction device is a wooden structure designed for controlled chest and abdomen compression. The device measures 48 inches in height and 33 inches in width. The structure consists of three side-by-side wooden boards, forming a rectangular shape from the top and side views (see Figures 1 and 2).

**Fig 1:**
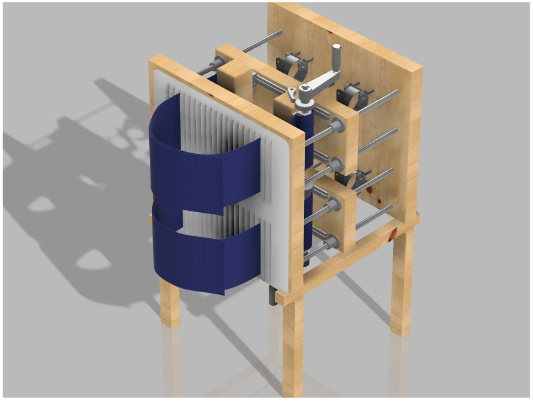
Side view of the 3D model of the chest wall and abdomen restriction device.

**Fig 2:**
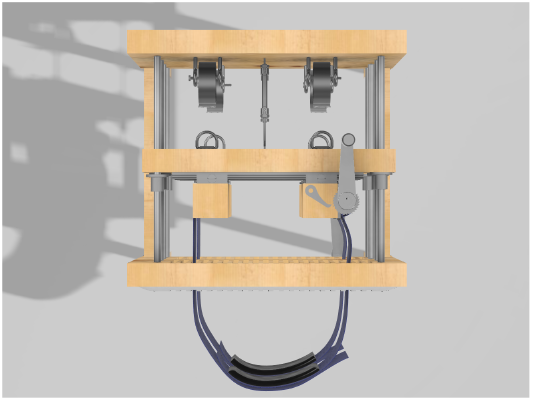
Top view of the 3D model of the chest wall and abdomen restriction device.

The device’s frame is built from a combination of precisely sized Compression Surface (front board), Rear Support Board (back board), Chest Restriction Board and Abdominal Restriction Board (middle boards), and the Structural Base, creating a strong and stable structure. This immobility is pivotal allowing the participate to be accurately compressed against the backboard.

The Compression Surface (front board) and Rear Support Board (back board) are identical, each measuring 24 inches wide, 22 inches high, and 2 inches thick. These dimensions were chosen to provide a sturdy backbone capable of supporting the device’s components and ensuring durability during use. The 24-inch width offers ample space for mounting other parts of the device, while the 22-inch height accommodates the average torso size of participants, ensuring the device can comfortably fit a range of body types.

The Compression Surface features a foam pad measuring approximately 22 by 22 inches and 0.5 inches thick. This padding was carefully sized to match the height of the Compression Surface and Rear Support Board, ensuring full coverage and comfort for the participant’s back. The 0.5-inch thickness and appropriate softness provide sufficient cushioning to distribute pressure evenly, reducing discomfort and preventing pressure points during use. Importantly, the material is not too soft, ensuring that the restriction applied by the device remains effective and is not compromised by the padding.

Between the Compression Surface and Rear Support Board are two middle boards: the Chest Restriction Board and Abdominal Restriction Board. These boards slide along four rods, which connect the Compression Surface and Rear Support Board. Each middle board is 22 inches wide, 8 inches high, and 2 inches thick, with a 3.3-inch vertical gap separating them. The 22-inch width aligns with the width of the Compression Surface and Rear Support Board, maintaining structural consistency. The 8-inch height provides enough surface area to cover the chest wall and abdomen of participants, as each board is responsible for holding the mechanism that restricts both the chest and abdomen separately.

The Structural Base of the device serves as the foundation, ensuring overall stability and balance. It consists of four identical legs, each 1.5 inches by 2 inches wide and standing 16.5 inches tall. These dimensions provide a sturdy base capable of supporting the device’s height and weight without tipping. The legs are connected by two horizontal wooden beams, each measuring 1.5 inches by 1.5 inches in width and 24 inches long. The horizontal beams distribute the load evenly across all four legs, preventing wobbling and enhancing the device’s durability during prolonged use.

#### 2) Rod Placement and Hooks

The rods connect the exterior wooden boards, creating a space of approximately 18 inches that allows the two middle boards to move freely. This spacing was chosen to provide enough room for placing the LVDTs (each 6.5 inches long) and for extending the springs to about 1.25 times their diameter (around 3 inches). More space might be needed depending on the LVDT being used and the diameter of the chosen springs. To support the springs, four metal hooks are positioned between the rods, as shown in Figures 2 and 5. Each hook is spaced 6 inches apart, with an additional 6-inch gap between the rods and the hooks.

#### 3) Strapping System

The blue vinyl strapping material, shown in Figure 3, is the device’s main component for applying restriction to the participant’s chest and abdomen, whether static or dynamic. Made from non-stretchable fabric, it prevents any unintended movement during use. Each strap is 3 feet long and 8 inches wide, providing enough material to fit individuals of various sizes. Once the participant is strapped in, the right side of the band overlaps the left side and is secured with Velcro. Then, the left band, connected to a metal handle and a ratchet gear mechanism, is tightened, pulling the right band along with it. The ratchet gear mechanism is shown in detail in Figure 4.

**Fig 3:**
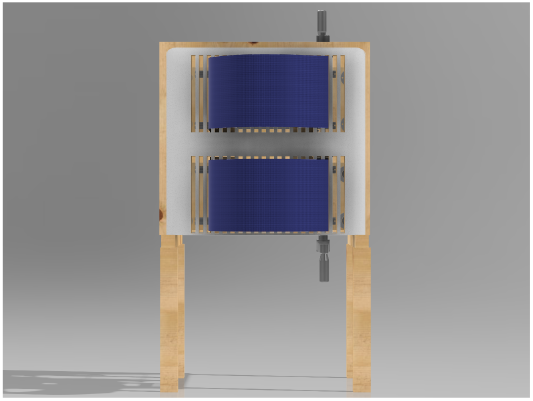
Front view of the 3D model of the chest wall and abdomen restriction device.

**Fig 4:**
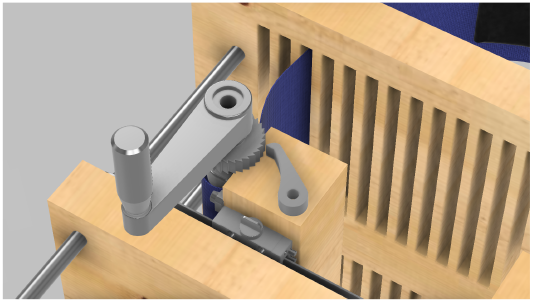
Close-up view of the ratchet gear mechanism used to adjust and secure the strapping system. This mechanism allows fine-tuning of the tension applied to the participant’s chest, ensuring precise control over compression.

#### 4) Dynamic Restriction and Constant Force Springs

The adjustable force springs that provide a constant force are essential to the device’s functionality. They are made from a flat strip of spring material coiled into a cylinder. Unlike traditional compression springs, these springs provide a steady force throughout their range of motion, reaching full strength after being stretched to 1.25 times their diameter. Once tensioned, they maintain a consistent force, allowing precise adjustment of the restriction applied to the participant’s chest and abdomen. The springs are mounted on the device’s backboard and connect to the tie-down rings via carabiners (not pictured). This setup, shown in Figure 5, enables the device to apply a consistent, controllable dynamic restriction as the participant breathes.

#### 5) Static Restriction Mechanism

For static restriction, the device uses four shaft collars, one on each rod. When these collars are positioned and tightened, the interior wooden walls are fixed in place, preventing any forward movement, as seen in Figure 6. This creates a static compression environment, allowing for studies where a constant chest or abdominal restriction is required.

**Fig 5:**
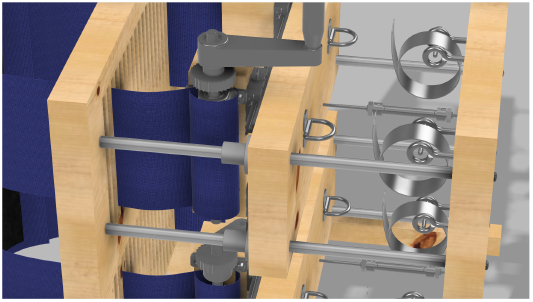
Close-up view of the dynamic load mechanism showing the constant force springs. Each spring is connected to a tie-down ring via a carabiner (not pictured).

**Fig 6:**
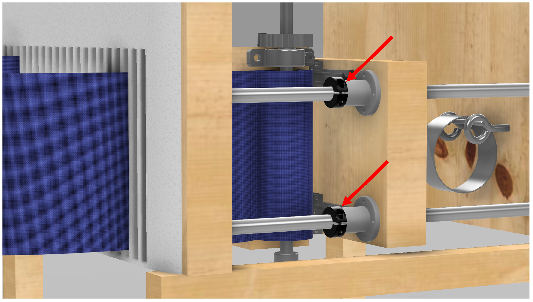
Close-up view of the static load mechanism. This image details the shaft collars positioned on the rods, which lock the interior wooden walls into place, creating a static restriction.

#### 6) Adjustable Width System

To ensure that the bands described in Section II-A3 fit snugly against the participant’s body, a mechanism was built into the middle boards to adjust the horizontal distance between the left and right bands for both the chest and abdomen separately.

Four wooden blocks (two for each middle board) move horizontally along metal rails mounted on the boards. These blocks, measuring 9 × 3.5 × 3.5 inches, slide on 19-inch rails attached to the interior walls (see Figure 7 for details). Slits cut into the front board, each 0.5 inches wide and spaced 1 inch apart, allow the bands to pass through and securely fit the participants’ body sizes.

**Fig 7:**
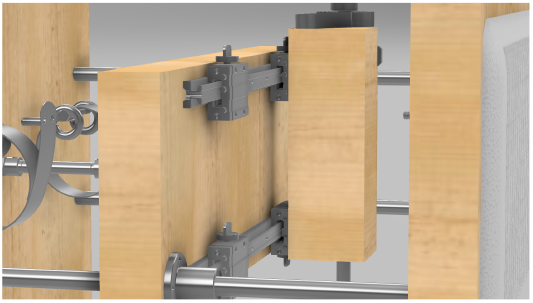
Close-up view of the adjustable width mechanism. The figure shows how the wooden boards slide along the metal rails, allowing controlled movement to adjust the width of the bands for a better fit.

#### 7) Pressure Sensors

The pressure sensors integrated into the strapping system are designed to accurately measure the pressure applied to the chest and abdomen, ensuring reproducibility and consistency in experimental conditions. These sensors (shown in black in Figure 8) function similarly to balloons with air pockets: both air pockets are inflated to the same volume (60 mL), and a sensor measures the internal pressure. Each air pocket, installed on the left band, measures 3.75 inches by 23 inches.

**Fig 8:**
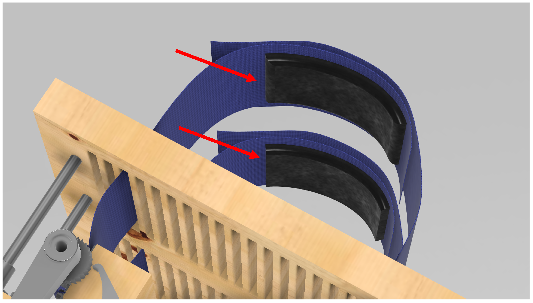
Close-up view of the integrated pressure sensors within the strapping system. These sensors are used to measure the pressure applied to the chest and abdomen, ensuring accurate and consistent experimental conditions.

### B. Participant Setup

The first step in setting up the participant in our procedure involves measuring the participant’s height and torso width. The participant then sits on an adjustable stool, initially measuring from the floor to the seat 16 inches and extending to a total height of 23 inches. We adjust the stool based on their chest and abdomen. The participant sits with their back against the front of the device, pressing onto the padded front wooden wall, and we use a metal gear handle to unwind enough vinyl strapping (loosen the straps) for a customized circumferential fit. We confirm the participant’s width and adjust the width of the bands to match using the adjustable width system (described in Section II-A6) and pass the bands through the corresponding slits on the front board. The goal is to align the straps parallel to the participant’s body.

Once the participant is properly seated, we use the vinyl straps as a preliminary check for chest and abdominal strapping, ensuring proper alignment before tightening. The lower vinyl strap fully encircles the abdomen, and the upper strap covers the chest once the proportions are confirmed. The participant sits upright, and we adjust the straps accordingly, securing the left vinyl strap onto the chest and the abdomen. The right strap is wrapped on top of the left strap and securely attached using Velcro.

For the dynamic condition, the chosen springs are mounted on the device’s backboard and connected to the tie-down rings using carabiners. For a static restriction, the springs are not used, and the two middle boards, which have rods running through them, are pulled back to their maximum position as restricted by the LVDT. Shaft collars are then attached to secure the wooden boards in place, establishing the static simulation setup.

#### 1) Load Calibration

Once fully strapped and positioned on the device, tension is applied using the metal gear. Adjusting the gear to different levels allows for varying pressure to be applied to the participant’s abdomen and chest. To ensure consistent pressure across participants, a calibration process is conducted. Participants are restricted by the bands and instructed to perform normal breathing for about a minute.

Using a live feed of data from the pressure sensors, the minimum pressure values, typically occurring at the final phase of expiration, are analyzed and used as the baseline pressure. If the baseline pressure is lower than the target pressure, the process is repeated: the restriction is tightened further, the participant performs normal breathing for another minute, and the baseline pressure is reassessed. This cycle continues until the baseline pressure matches the target pressure.

### C. Experimental Design Validation

#### 1) RQ1: Breathing Restriction - Static and Dynamic Design Validation

To validate the capability of the device to enable both static and dynamic restriction within the same design, an experiment was conducted to evaluate how the device affects breathing under these conditions. Four males and two females (mean age 24.0 ± 3.4 years), all in good health, volunteered to participate and completed the entire study. The study procedures were approved by the Institutional Review Board of the University of Florida and adhered to the standards set by the Declaration of Helsinki.

The study protocol included three conditions: baseline (without restriction), static restriction, and dynamic restriction (with a load of 45 kg). Participants performed the tasks in a fixed order: baseline condition first, followed by the static restriction and the dynamic restriction. For each condition, pulmonary function testing was conducted to measure forced expiratory volume in one second (FEV1) and forced vital capacity (FVC). At least three tests were performed per condition, ensuring they met the acceptability criteria of the American Thoracic Society and European Respiratory Society, with the highest values recorded. Additionally, after each pulmonary function test, participants were asked to breathe normally for 5 minutes. Respiratory data, including airflow and mouth pressure, were recorded using a pneumotach connected to a mouthpiece and a two-way non-rebreathing valve (Hans Rudolph).

These measurements provided quantitative metrics to confirm differences between static and dynamic conditions, as indicated by LVDT displacement and respiratory changes. The data were tested for equal variance and normality, revealing non-normal distribution. Consequently, the Kruskal-Wallis test, a non-parametric alternative to ANOVA, was used to analyze differences between conditions, ensuring the validity of the statistical evaluation.

#### 2) RQ2: Reproducible and Adjustable Loads - Pressure Variability Validation

To evaluate the reproducibility and adjustability of the loads applied to the chest and abdomen, a second experiment was conducted focusing on pressure variability. The built-in pressure sensors of the device were connected to a data acquisition system, enabling real-time monitoring and recording of pressure. One participant was strapped in and instructed to breathe for 1 minute, then fully released. This process was repeated three times for each condition (static and dynamic), with pressure data from both the abdomen and chest recorded during each trial.

The pressure signals were analyzed using MATLAB, where the trough of each respiratory oscillation was selected as the fiducial point to measure pressure, as shown in Figure 9. These troughs were identified as the local minima in the pressure signal and used to define a baseline pressure, representing the minimum pressure acting on the participant during each breath. The baseline pressure *P*_baseline_(*i*) for the *i*-th breath was calculated as follows:

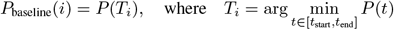

**Fig 9:**
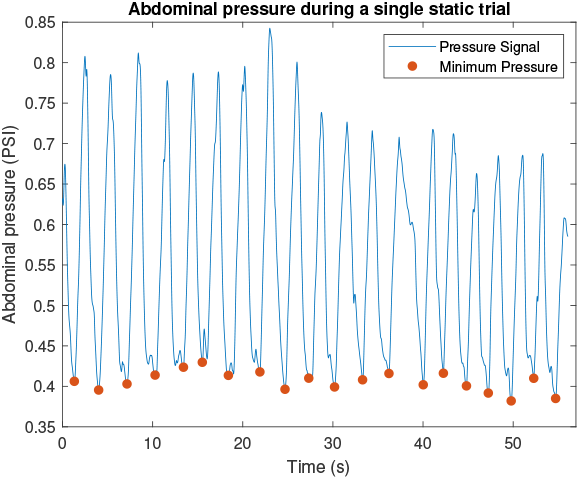
Example of how abdominal pressure varies within a single trial. Dots show the baseline pressure.

Here, *T*_*i*_ is the time point where the *i*-th local minimum occurs within the time interval [*t*_start_, *t*_end_], representing one respiratory cycle. The pressure value *P*_baseline_(*i*) quantifies the minimum pressure for the *i*-th breath, providing an objective measure of load variability and reproducibility.

This experiment demonstrated the ability of the device to apply consistent and adjustable loads across trials, confirming its reproducibility and flexibility. The use of fiducial points ensures precise calibration and measurement, validating the device’s performance in achieving adjustable and reproducible load conditions.

## III. Results

### A. RQ1: Static and Dynamic Restriction Validation

The ability of the device to achieve both static and dynamic restriction was validated through respiratory metrics and displacement measurements. Figure 10 summarizes the results for various respiratory metrics across the baseline, static, and dynamic conditions.

**Fig 10:**
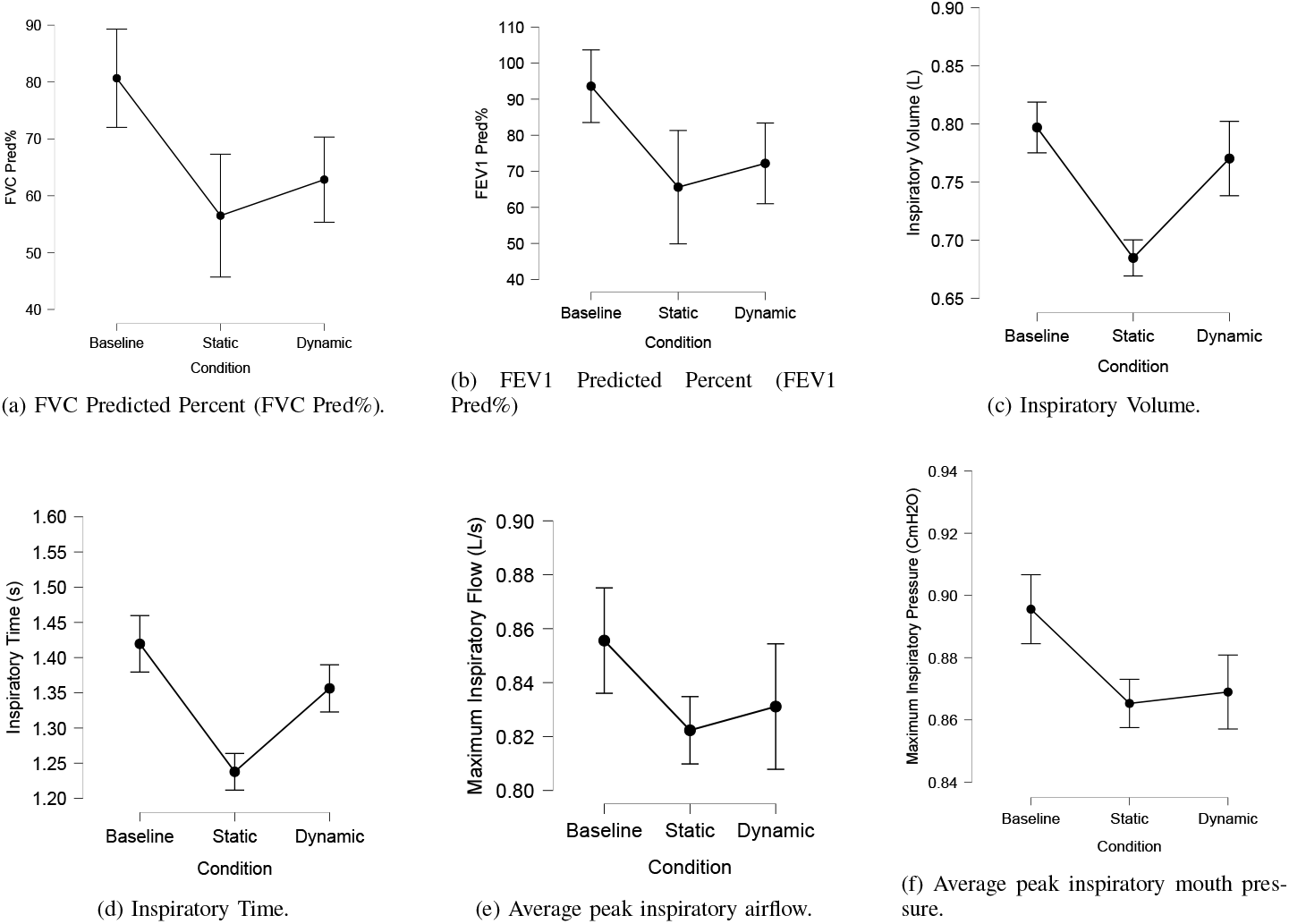
Summary of various respiratory metrics across different conditions: baseline, static, and dynamic.

The Kruskal-Wallis test revealed statistically significant differences in FEV1 Predicted % (Figure 10a) among the three conditions (Baseline, Static, and Dynamic), with a p-value of 0.011. Dunn’s Post Hoc Comparisons highlighted significant differences between Baseline and Static (p = 0.004) and between Baseline and Dynamic (p = 0.026), with no significant difference between Static and Dynamic conditions (p = 0.524). Similarly, for FVC Predicted % (Figure 10b), significant differences were observed between Baseline and both Static (p = 0.002) and Dynamic (p = 0.017) conditions, while Static and Dynamic did not differ significantly (p = 0.448).

Additional metrics, including inspiratory volume, inspiratory time, peak inspiratory airflow, and peak inspiratory pressure, also demonstrated significant differences across conditions (p < 0.001). Inspiratory volume (Figure 10c) and inspiratory time (Figure 10d) varied significantly between Baseline and both Static and Dynamic conditions, confirming the ability to simulate different restriction levels. Maximum inspiratory airflow and pressure (Figures 10e and 10f) further emphasized the distinctions between static and dynamic setups, with lower values observed during the dynamic condition.

Figure 11 highlights the displacement of the LVDTs during one minute for a single participant. During the static condition, minimal movement was observed, with displacements close to zero (approximately 5 × 10^−3^ cm), while the dynamic condition demonstrated clear movement of both the chest and abdomen, with displacements reaching approximately 0.5 cm. These results validate the device’s ability to achieve distinct static and dynamic conditions.

**Fig 11:**
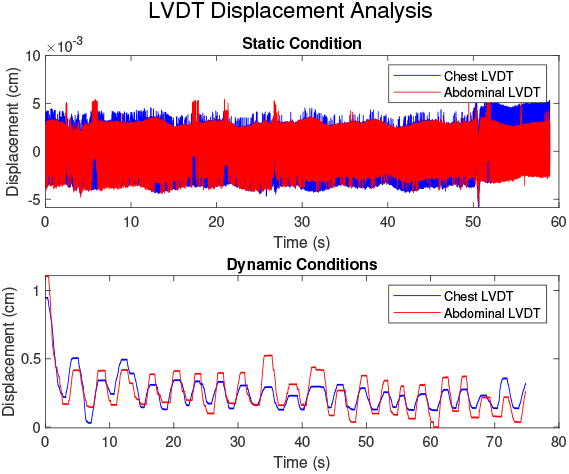
Displacement of the LVDT across static and dynamic conditions for a single participant over a one-minute period. During the static condition, minimal movement is observed, with displacements close to zero (approximately 5 × 10*-*3 cm). In contrast, the dynamic condition demonstrates clear movement of both the chest and abdomen, with displacements reaching approximately 0.5 cm.

### B. RQ2: Reproducibility and Adjustability of Load

The reproducibility and adjustability of the device were validated by analyzing pressure variability across trials for both static and dynamic conditions. Pressure data were recorded for the chest and abdomen, and the results are summarized in Tables I and II.

**TABLE I:** Static Condition for Baseline Pressure.

|  |  | Mean | Std. Deviation |
| --- | --- | --- | --- |
| Minimum Chest Pressure (PSI) | 1 | 0.471 | 0.024 |
|  | 2 | 0.461 | 0.027 |
|  | 3 | 0.494 | 0.032 |
| Minimum Abdominal Pressure (PSI) | 1 | 0.430 | 0.011 |
|  | 2 | 0.406 | 0.012 |
|  | 3 | 0.392 | 0.013 |

**TABLE II:** Dynamic Condition for Baseline Pressure.

|  |  | Mean | Std. Deviation |
| --- | --- | --- | --- |
| Minimum Chest Pressure (PSI) | 1 | 0.441 | 0.017 |
|  | 2 | 0.400 | 0.057 |
|  | 3 | 0.386 | 0.038 |
| Minimum Abdominal Pressure (PSI) | 1 | 0.390 | 0.012 |
|  | 2 | 0.404 | 0.076 |
|  | 3 | 0.393 | 0.037 |

In the static condition (Table I), chest pressure showed consistent mean values across trials: 0.471 PSI (SD = 0.024), 0.461 PSI (SD = 0.027), and 0.494 PSI (SD = 0.032). Similarly, minimum abdominal pressures were consistent, with means of 0.430 PSI (SD = 0.011), 0.406 PSI (SD = 0.012), and 0.392 PSI (SD = 0.013).

In the dynamic condition (Table II), the minimum chest pressure exhibited a decrease across trials, with mean values of 0.541 PSI (SD = 0.017) in the first trial, 0.400 PSI (SD = 0.057) in the second, and 0.386 PSI (SD = 0.038) in the third. The minimum abdominal pressure also showed slight variations, with mean pressures of 0.390 PSI (SD = 0.012), 0.404 PSI (SD = 0.076), and 0.393 PSI (SD = 0.037).

These results demonstrate the device’s capability to apply reproducible and adjustable loads, supported by consistent pressure measurements across trials for both static and dynamic conditions.

## IV. Discussion

### A. RQ1: Static and Dynamic Restriction Validation

The findings demonstrate that our CWR device effectively modulates respiratory function, influencing both lung function measures and respiratory mechanics across static and dynamic conditions. The significant differences in FEV1 Predicted% and FVC Predicted% between baseline and the restricted conditions (static and dynamic) indicate that the device successfully induces a restriction [30].

Inspiratory volume, inspiratory time, and maximum airflow showed statistically significant differences between baseline and each restricted condition, further validating the effectiveness of the device. These findings align with previous research on the work of breathing under load and compensatory mechanisms [31], [32], [33]. The equation that describes work of breathing using airflow, first defined by [34] and later updated by [33],

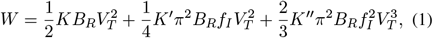

shows that the work of breathing is influenced by tidal volume (*V*_*T*_), breathing rate (*B*_*R*_), and inspiratory frequency (*f*_*I*_). Subjects adapt their breathing patterns under load by reducing inspiratory time and maximum airflow, resulting in shallower and longer breaths. This adaptation decreases the work of breathing by reducing instantaneous breathing frequency, consistent with the behavior observed under restricted conditions.

The data from the LVDTs further confirm the effectiveness of the static restriction, with virtually zero movement recorded during the static trials, validating the ability of the device to enforce a rigid, non-movable condition. In contrast, the dynamic condition allowed for measurable displacement of both the chest and abdomen, highlighting the device’s ability to simulate dynamic breathing scenarios. These results demonstrate that the CWR device achieves distinct and quantifiable static and dynamic restrictions.

### B. RQ2: Reproducibility and Adjustability of Load

The experiment designed to measure the variability of pressure exerted on the chest and abdomen showed that in the static condition, both chest and abdominal pressures were highly consistent across repeated trials. This consistency validates the reproducibility of the device’s load application. The data also confirm that the pressure levels can be adjusted and maintained precisely, supporting the adjustability of the device. In the dynamic condition, the pressure and displacement data indicate the device’s ability to allow separate movements of the chest and abdomen, offering a flexible platform to study respiratory mechanics difference between chest and abdominal breathing [35], [36]. Further, the combined data from the LVDTs and pressure sensors enable future research on the quantification of breathing patterns, muscle group activation and respiratory effort.

### C. Future Directions

Future research should address the effectiveness of the device across various body types. Although the design includes features to accommodate different body shapes and sizes, extreme body types may challenge full conformity to the device. Enhancing customization options for the device remains critical to improving its adaptability and effectiveness across a broader user base. Additional studies could refine the device to better address the needs of individuals with extreme body sizes.

Further research should also explore the use of the device in populations with respiratory diseases such as asthma. Special attention must be given to avoid triggering bronchoconstriction, and load levels should be carefully adjusted for such populations. Investigating device modifications tailored for individuals with compromised pulmonary function, such as adjustable spring resistance or softer band materials, could expand the device’s applications and improve its safety and efficacy.

Finally, the potential applications of this device extend beyond respiratory function. Future studies could evaluate its utility in assessing cognitive function, muscle fatigue, or blood circulation under restricted conditions, broadening the impact of this novel technology.

## V. Conclusion

Our CWR device offers a novel design to study the impact of chest wall impairment on breathing and cognition. It is an improvement on earlier devices, offering improved precision and versatility while allowing for accurate quantification of the load applied to the chest wall and abdomen. Thus, it provides researchers with controlled conditions for exploring the impacts of static and dynamic load-based restrictions on physiological and cognitive processes and modeling breathing dynamics under load [33]. Our validation studies have demonstrated significant experimental manipulation in the breathing process. Additional use cases for this device include respiratory training, assisting patients with respiratory conditions such as neuromuscular diseases [37], [38], and reducing the effort required to breathe [39], [40].

## Data Availability

All data produced in the present study are available upon reasonable request to the authors.

## Appendix A Overview of the design

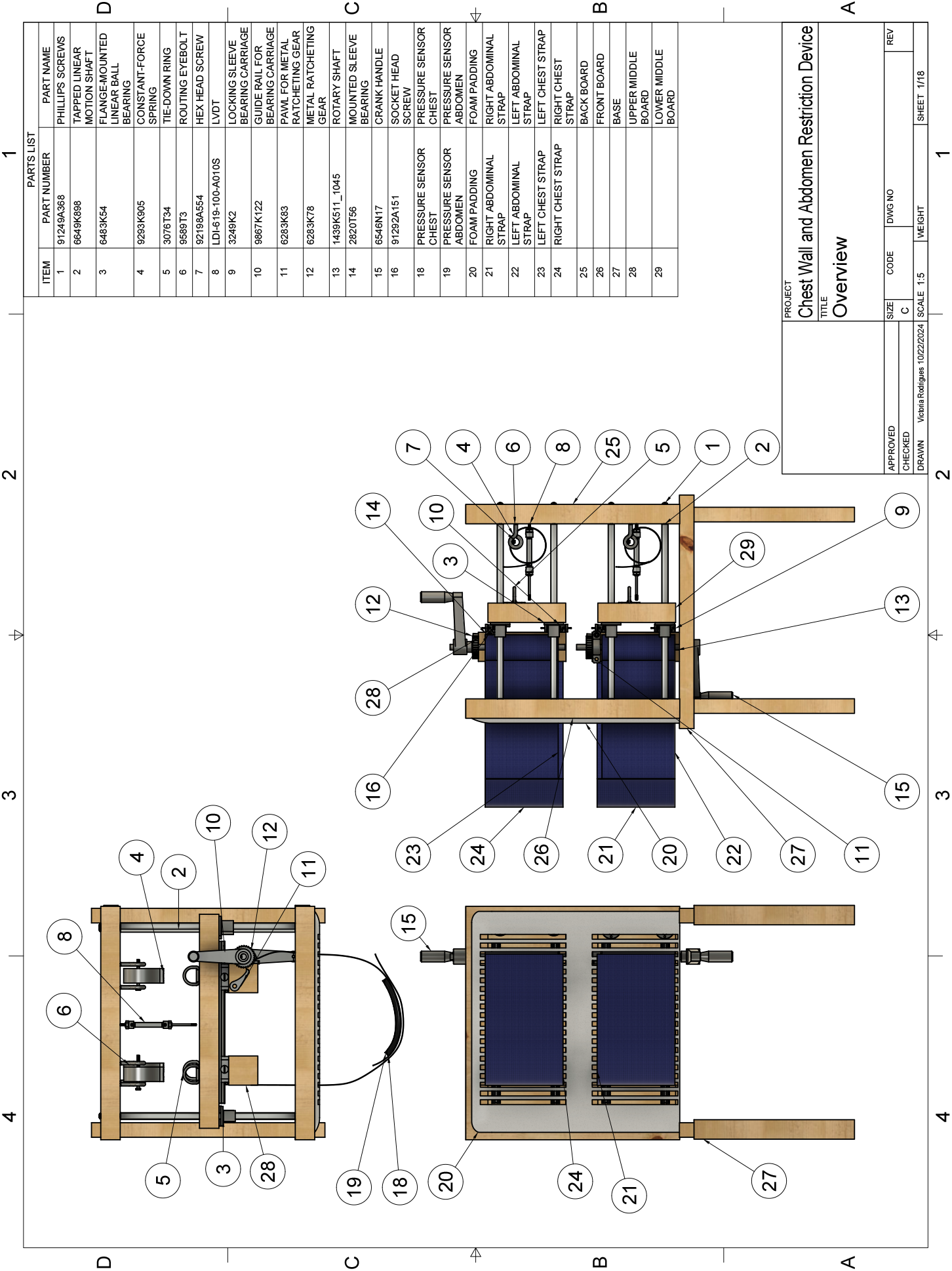

## Appendix B Dimensions

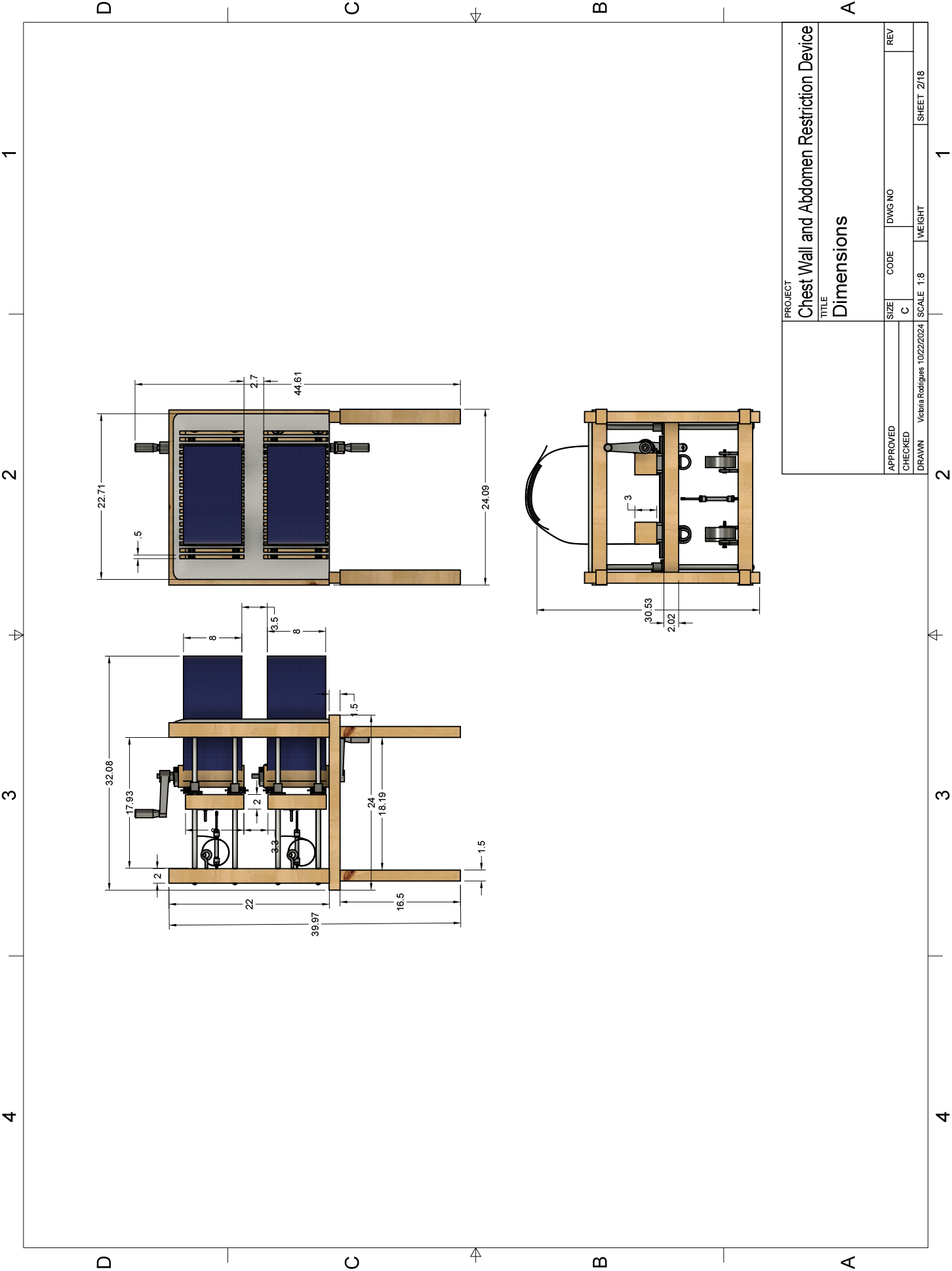

## Appendix C Structural Base

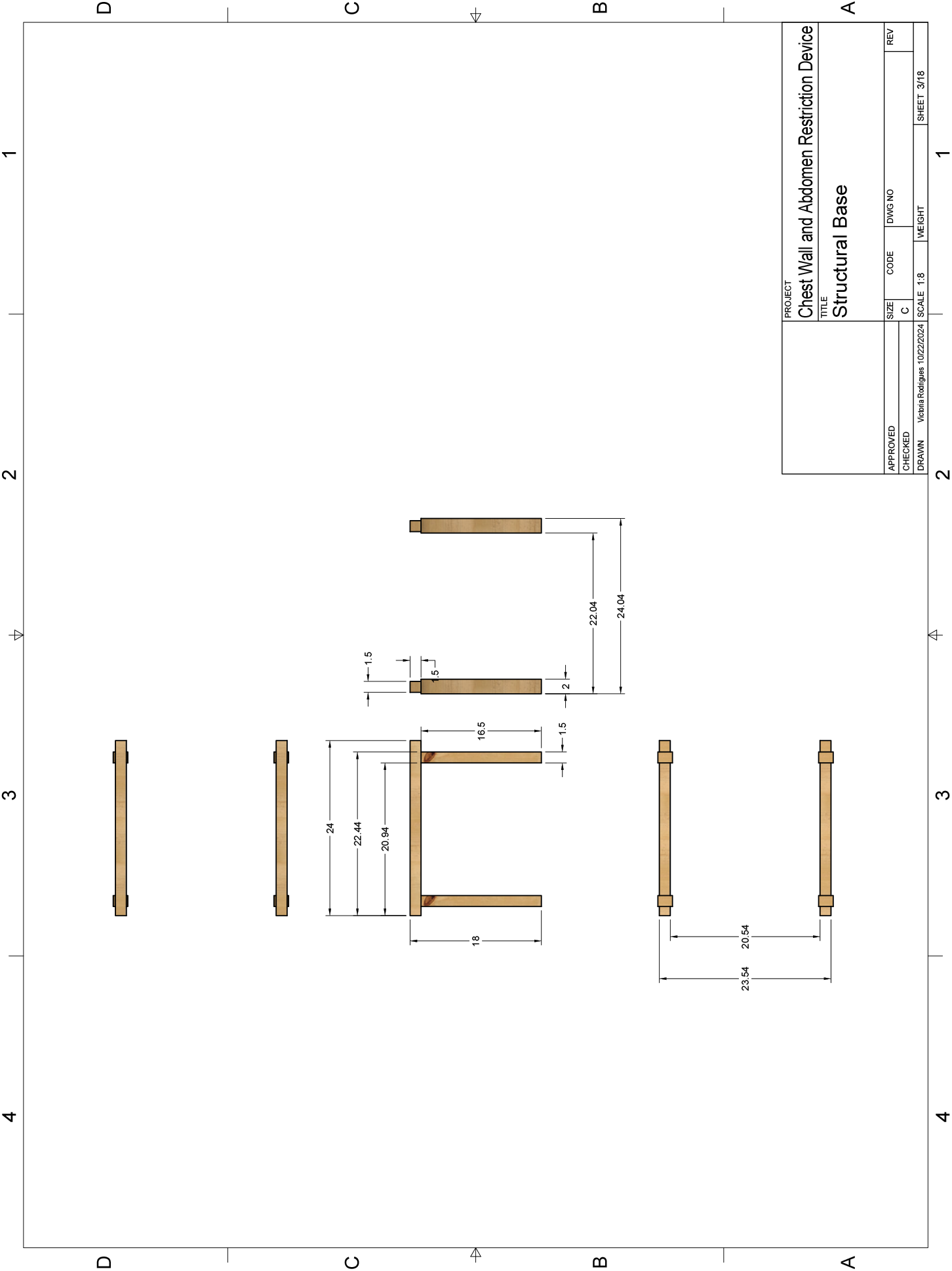

## Appendix D Compression Surface

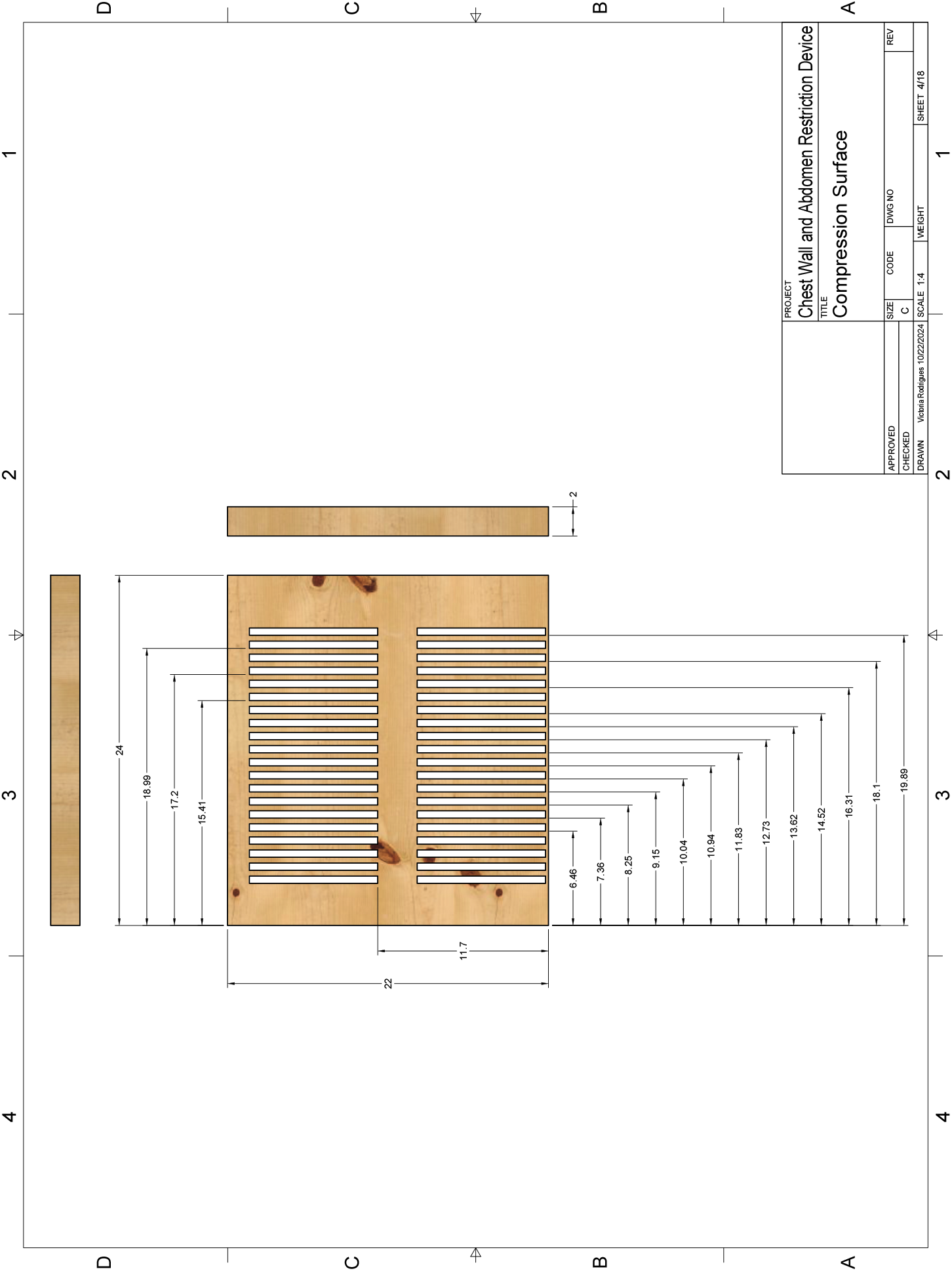

## Appendix E Rear Support Board

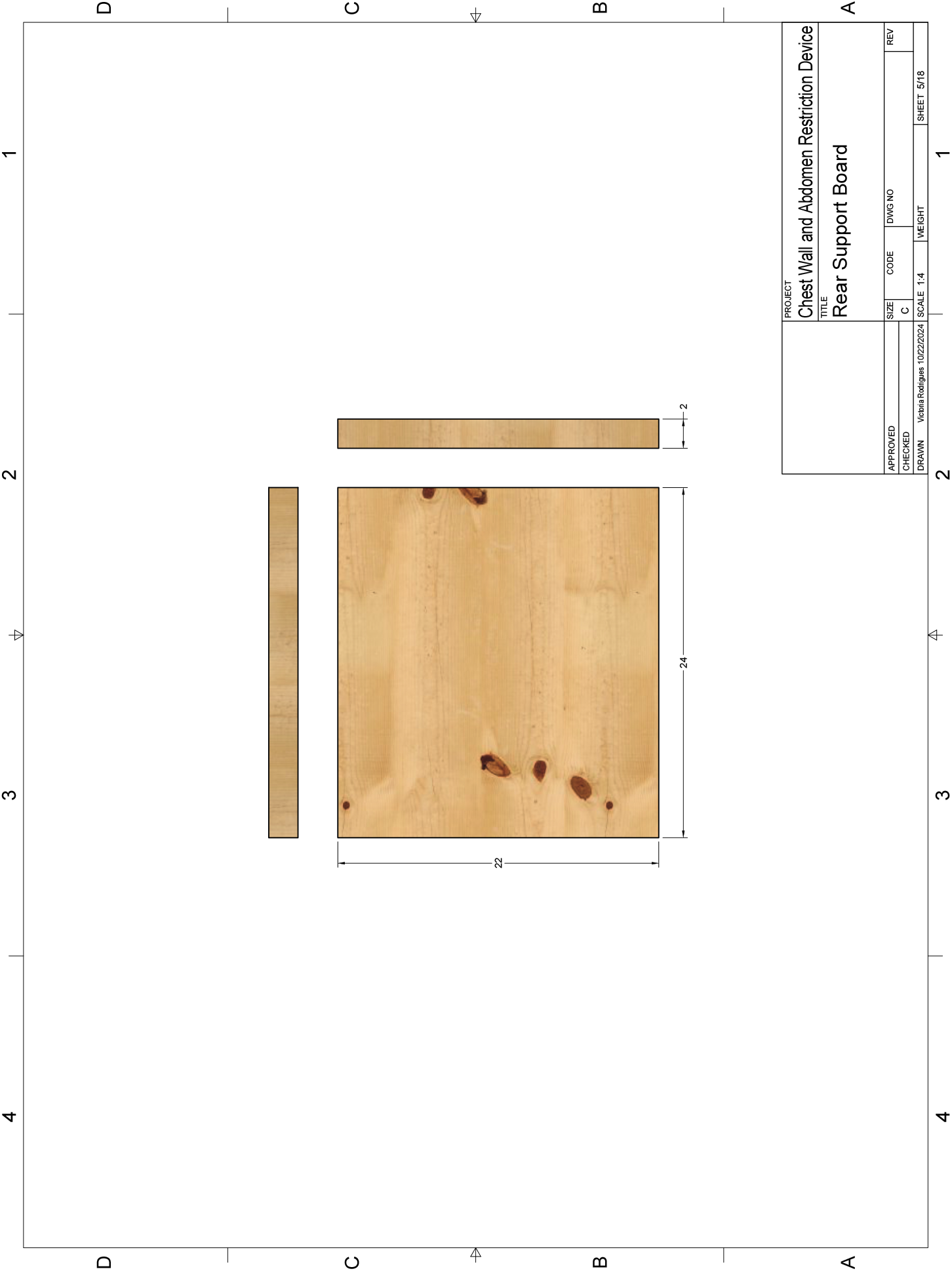

## Appendix F Chest and Abdominal Restriction Boards

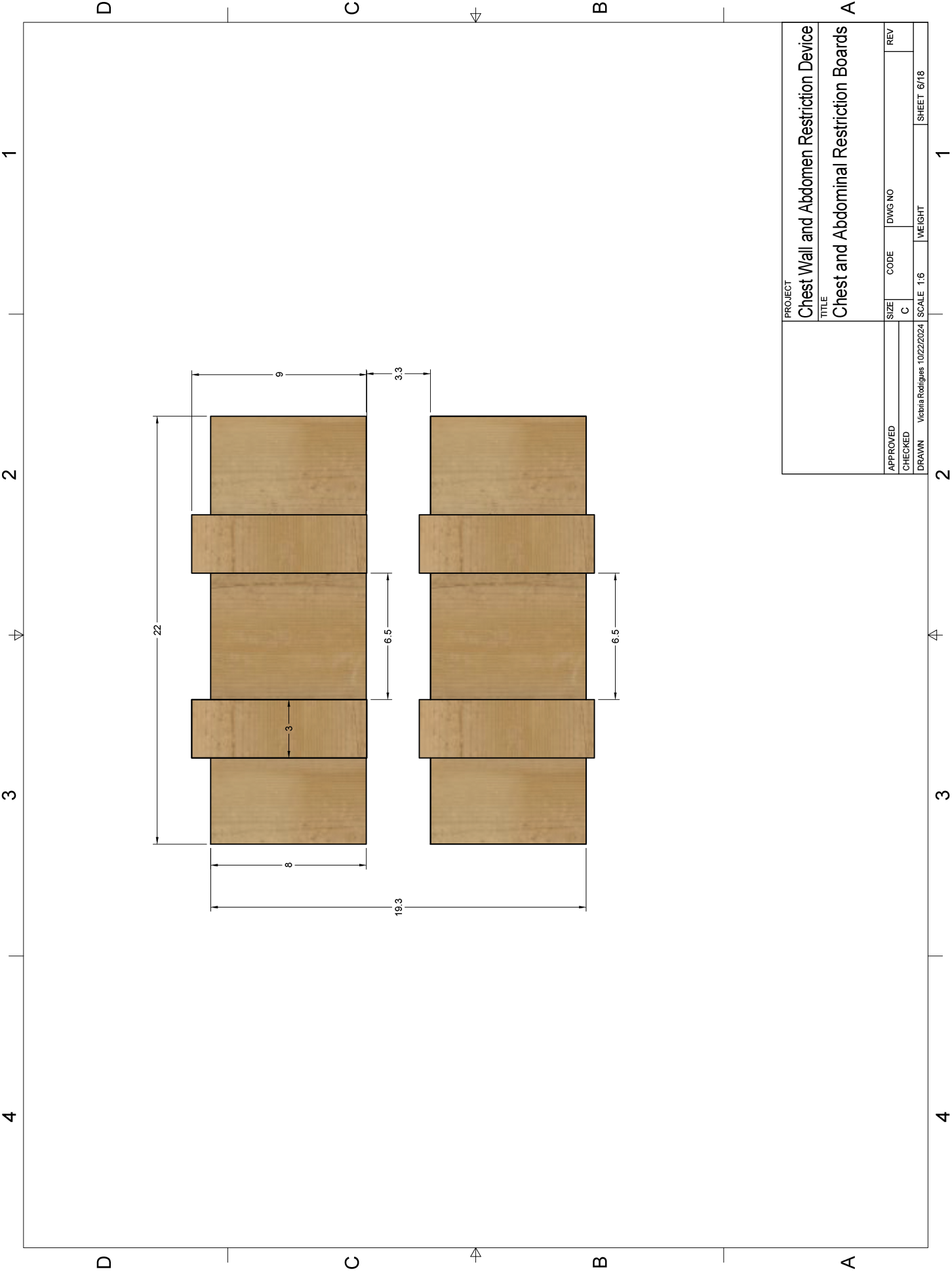

## Appendix G Foam Padding

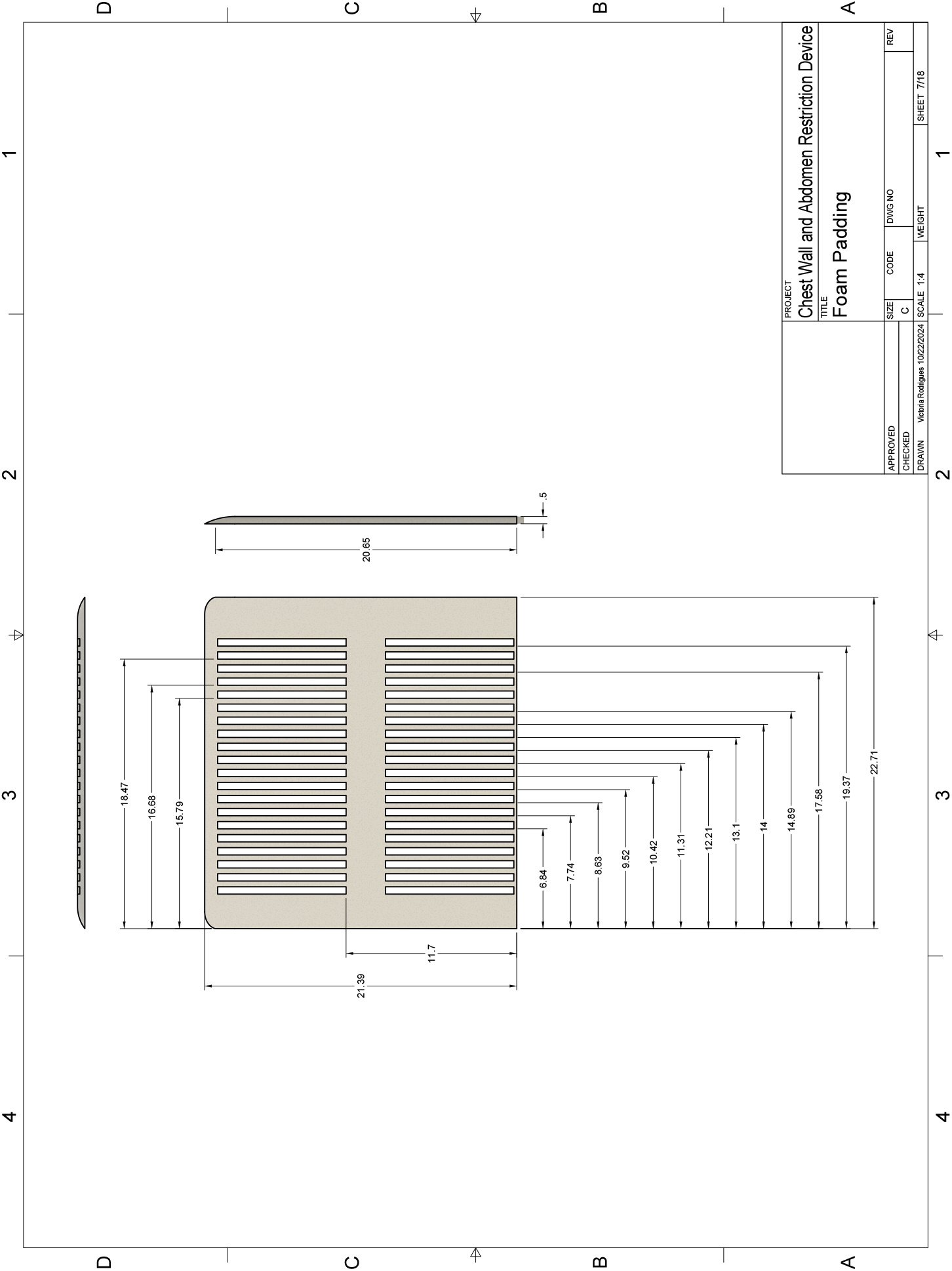

## Appendix H Flanged-Mounted Linear Ball Bearing

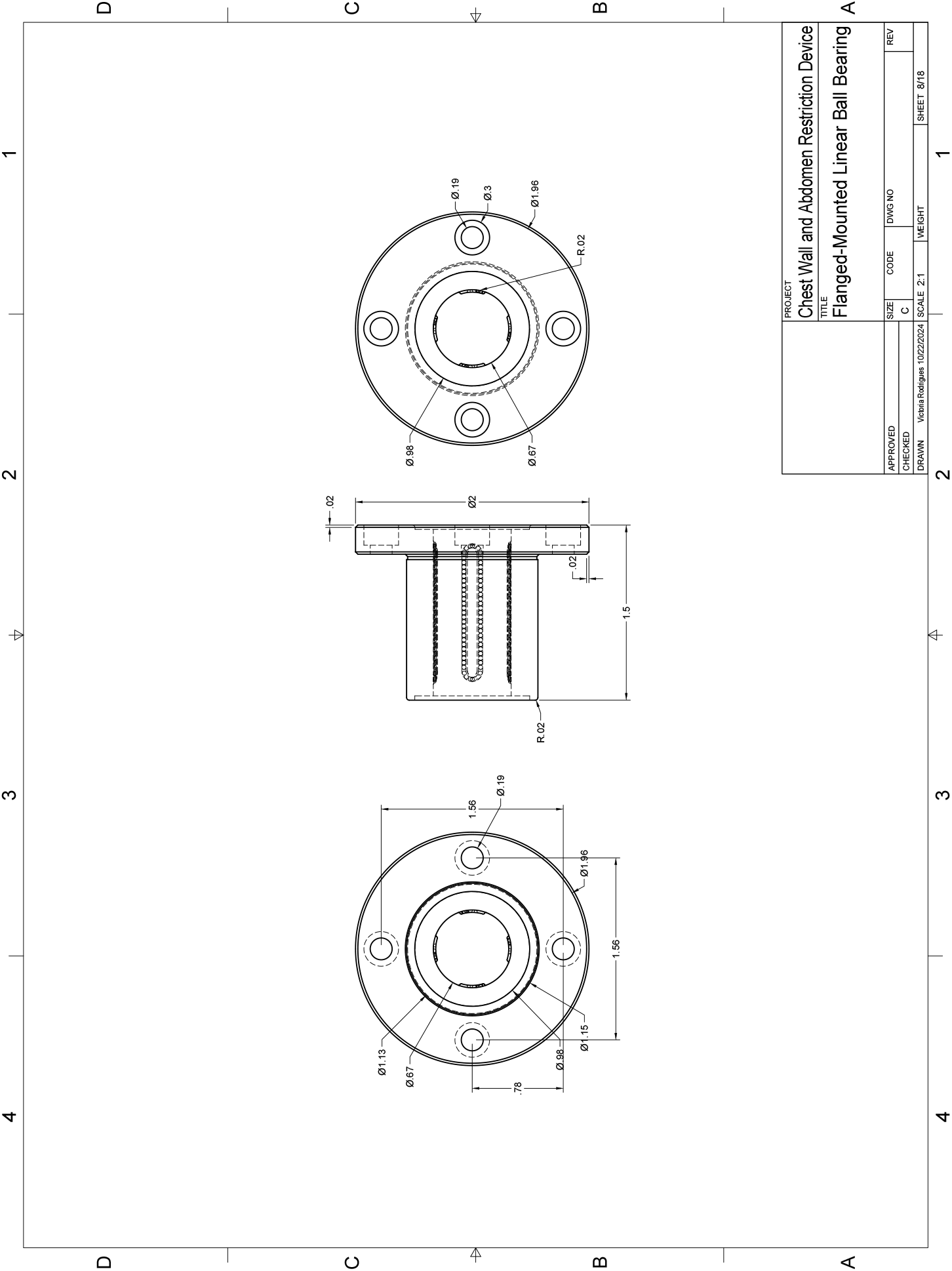

## Appendix I Constant-Force Spring

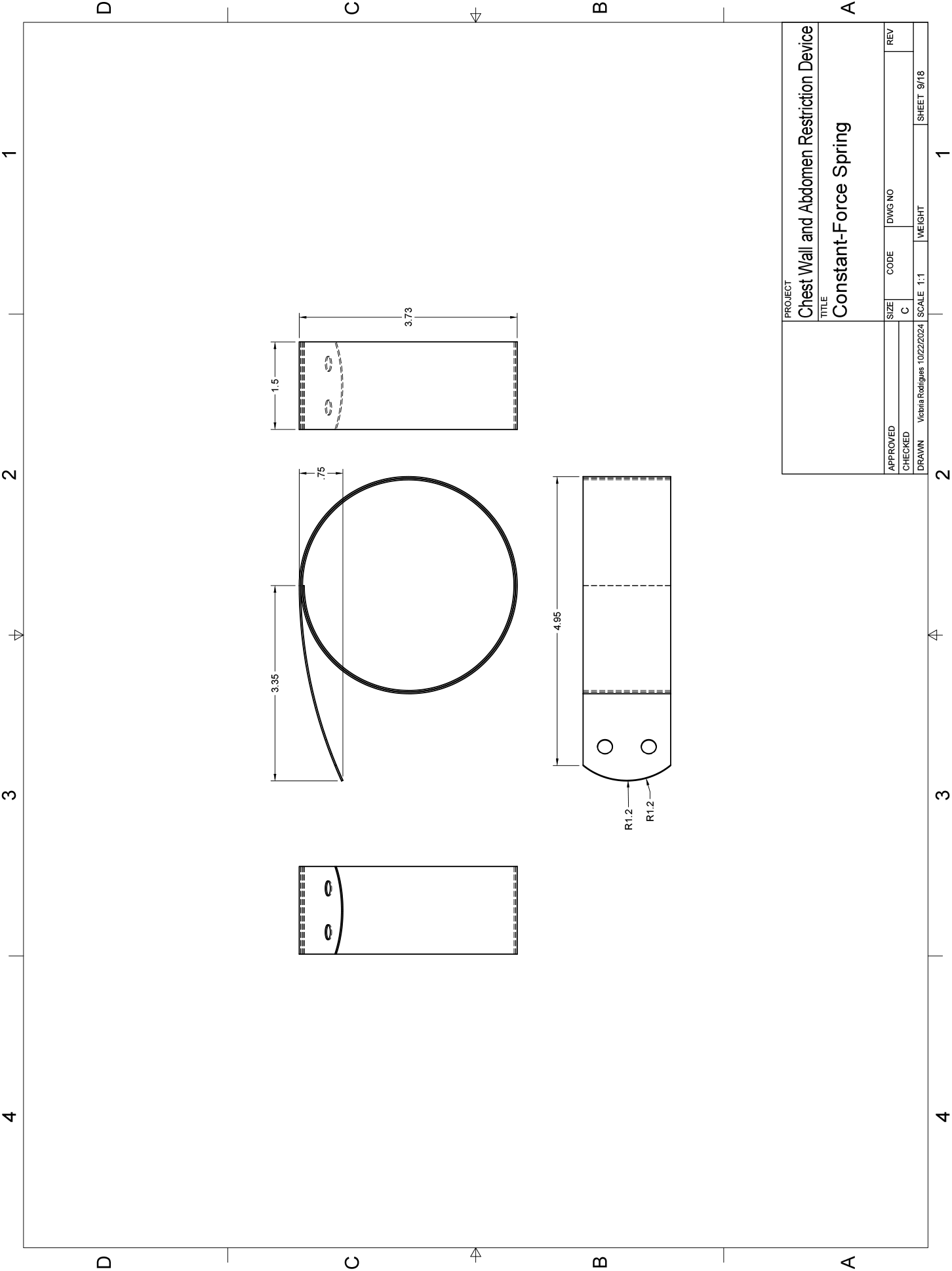

## Appendix J Tie-Down Ring

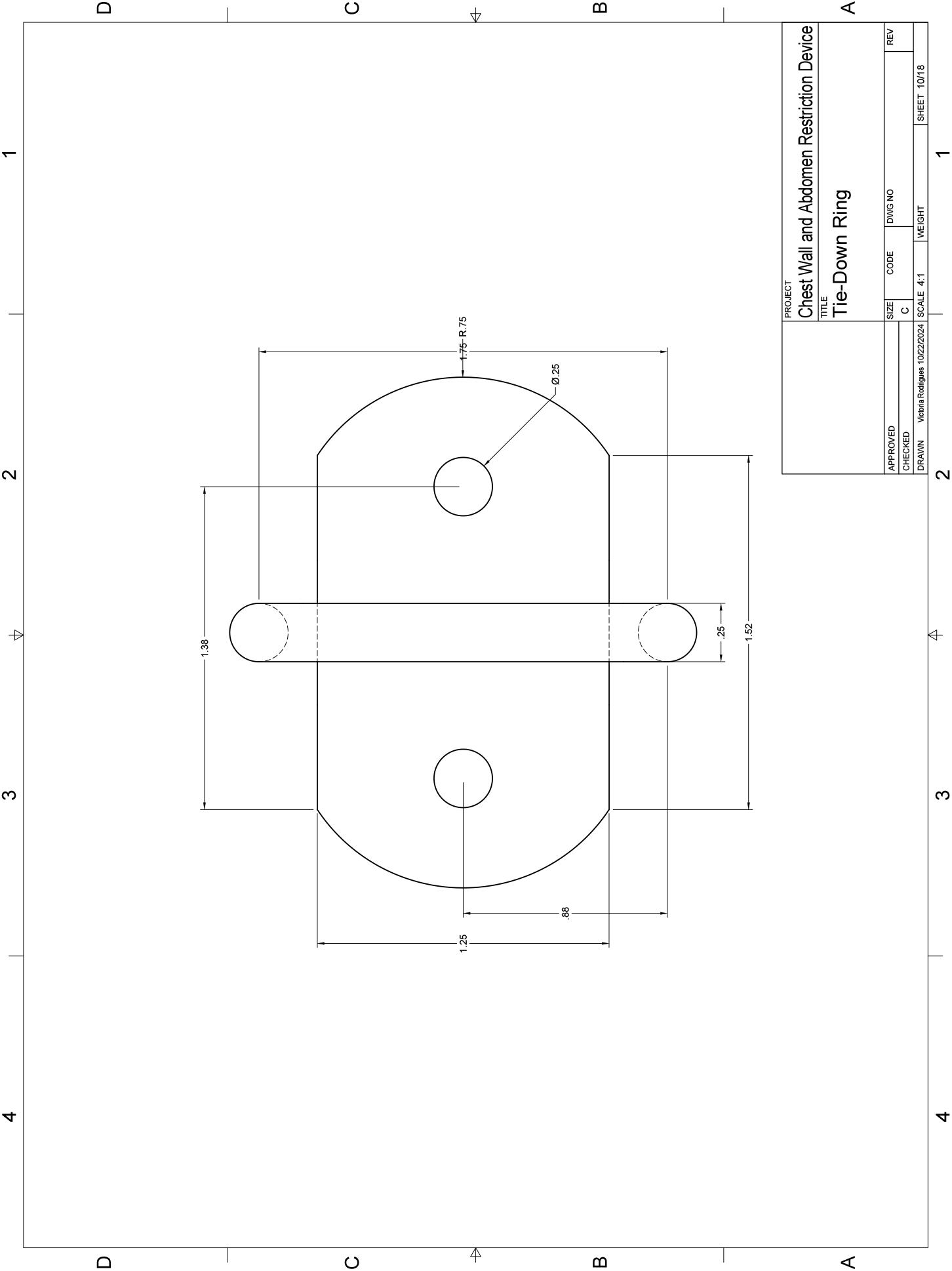

## Appendix K Locking Sleeve Bearing Carriage

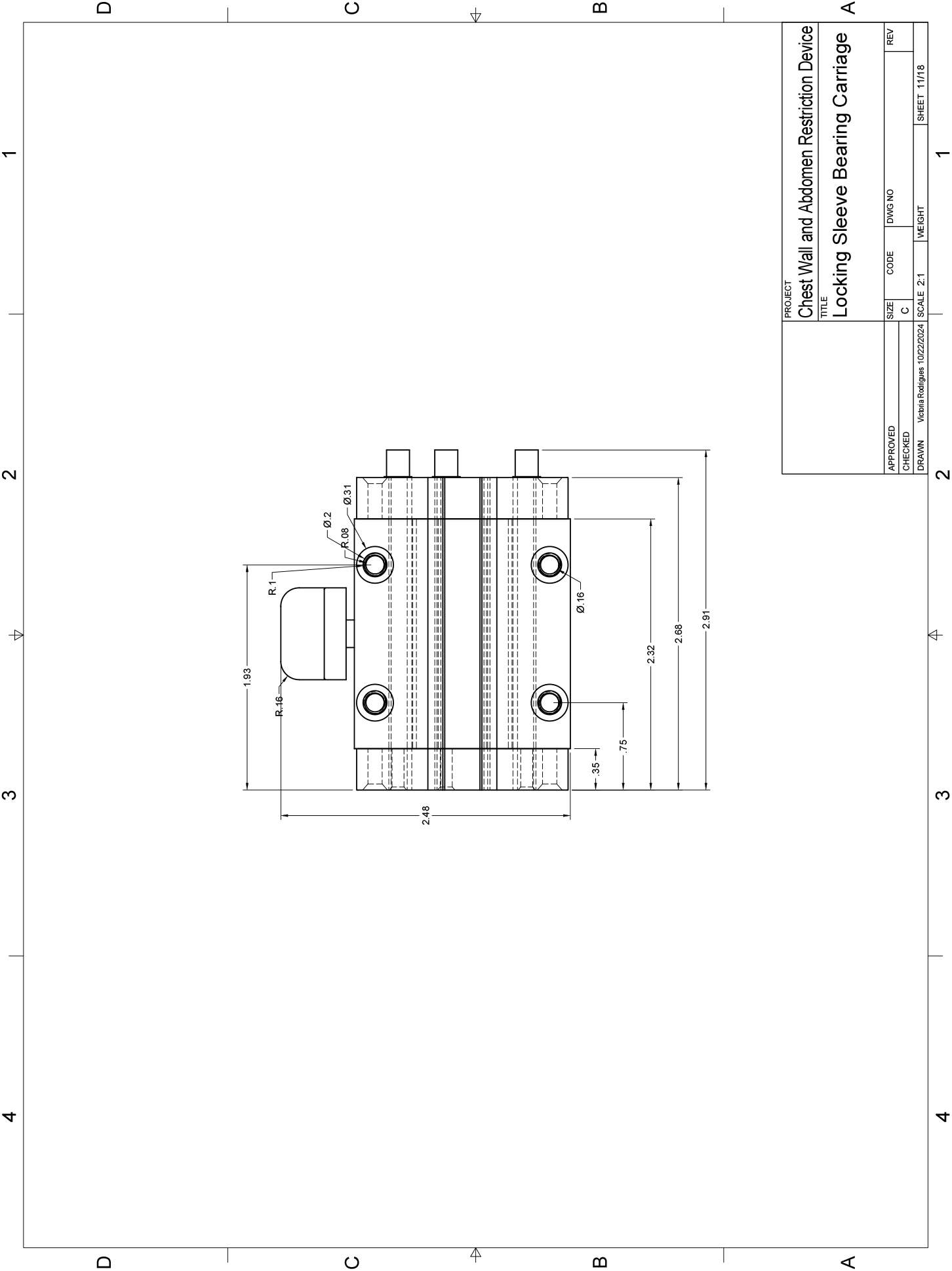

## Appendix L Guide Rail for Bearing Carriage

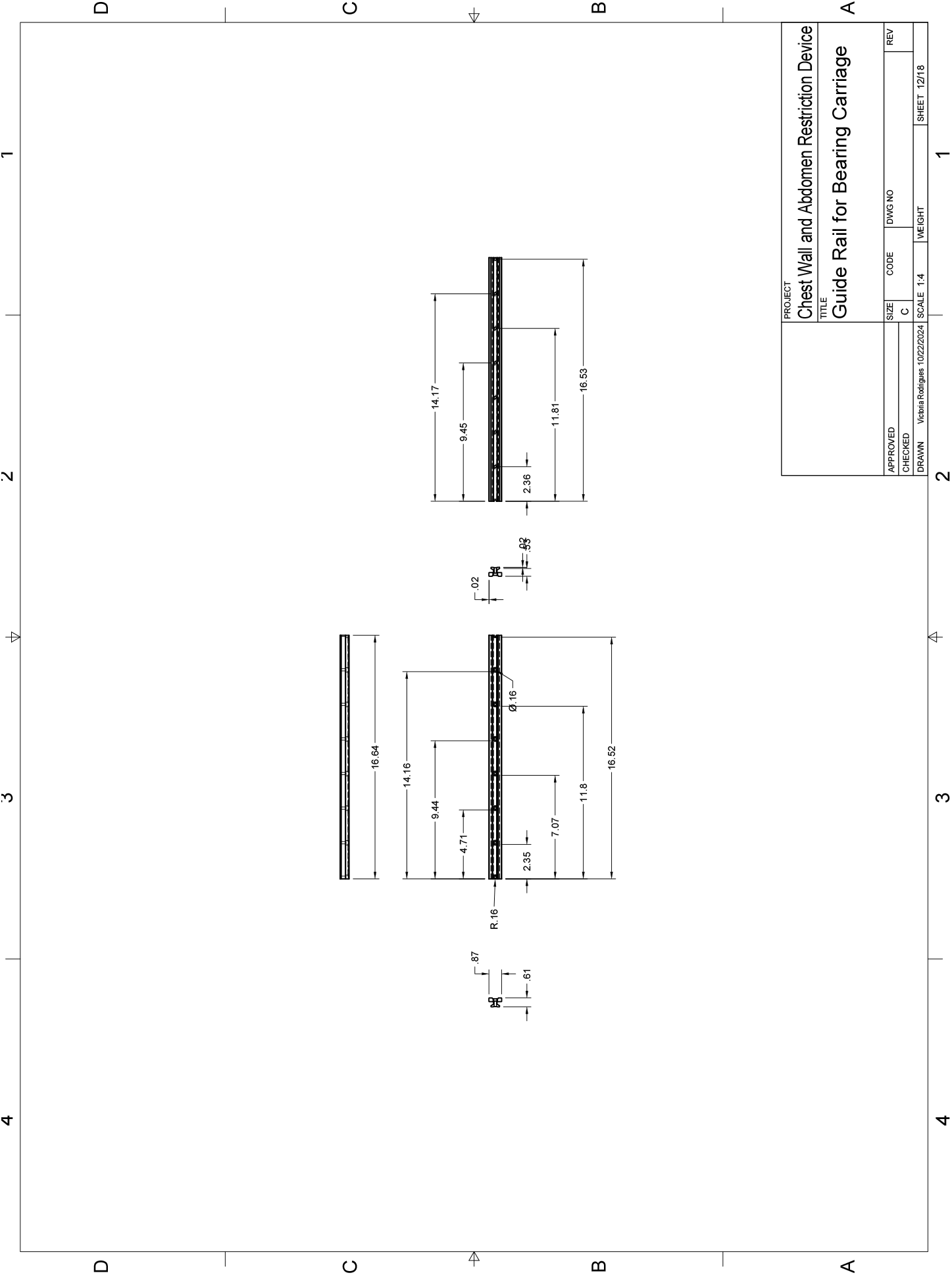

## Appendix M Pawl for Metal Ratcheting Gear

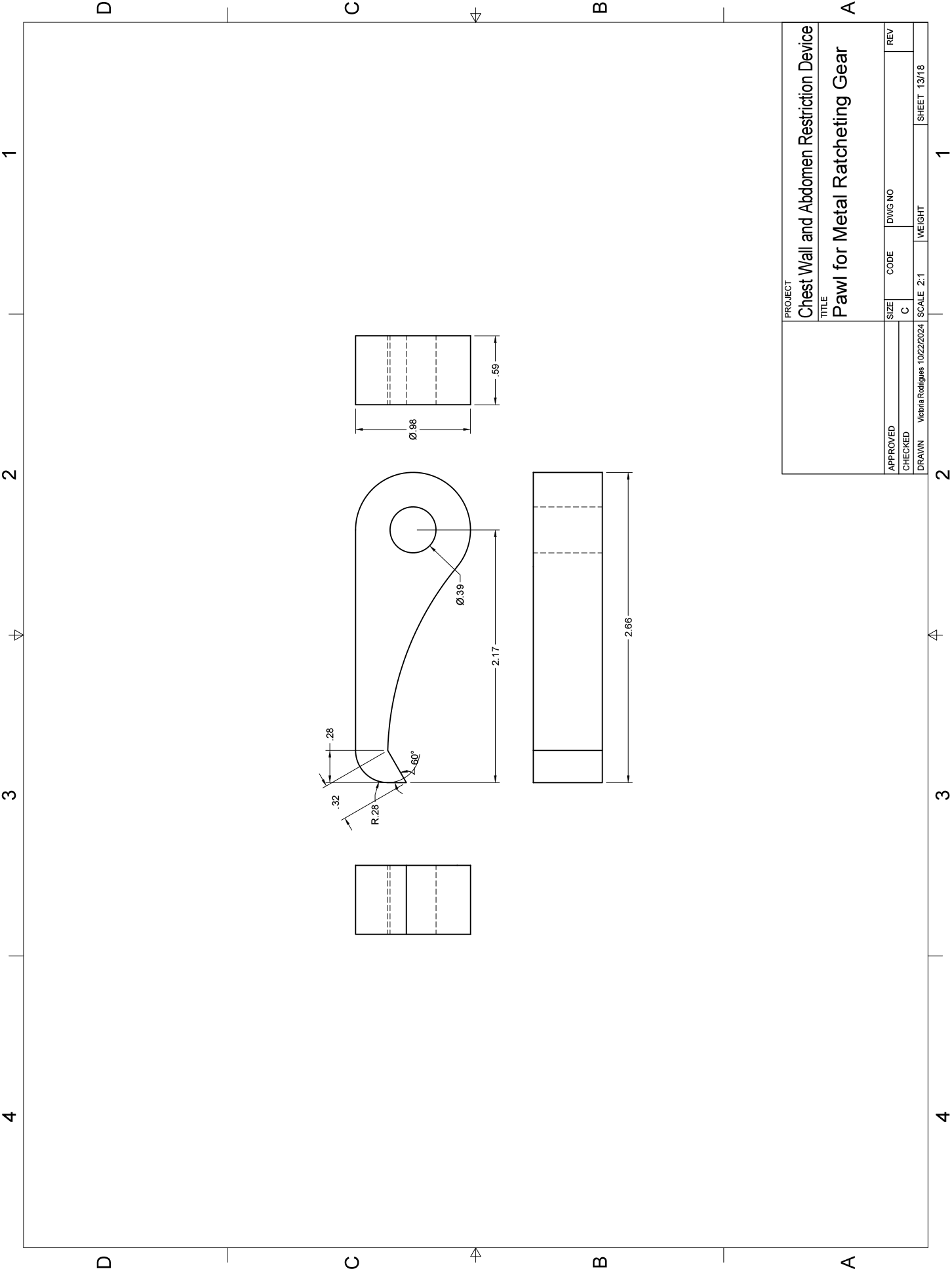

## Appendix N Metal Ratcheting Gear

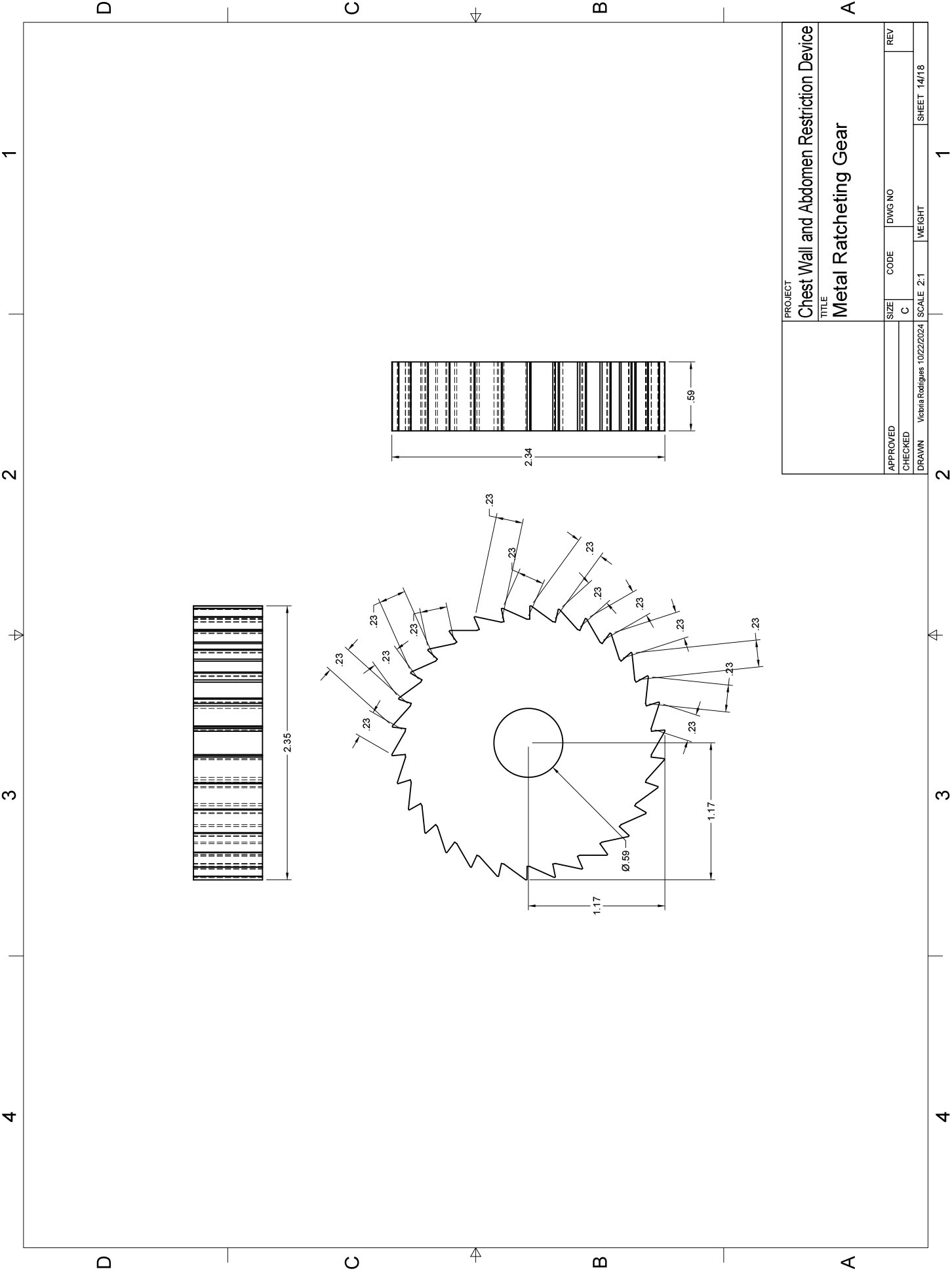

## Appendix O Rotary Shaft

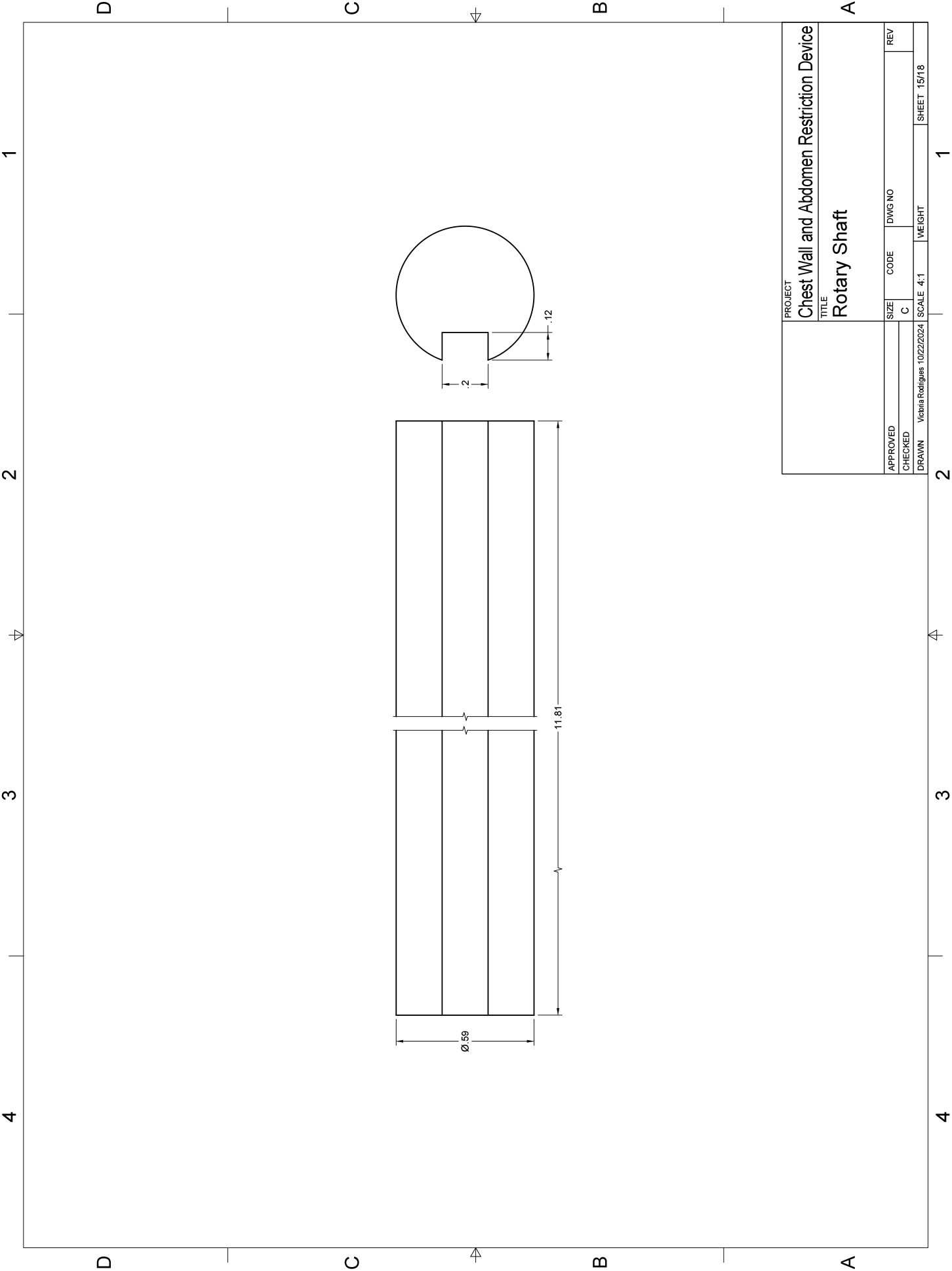

## Appendix P Mounted Sleeve Bearing

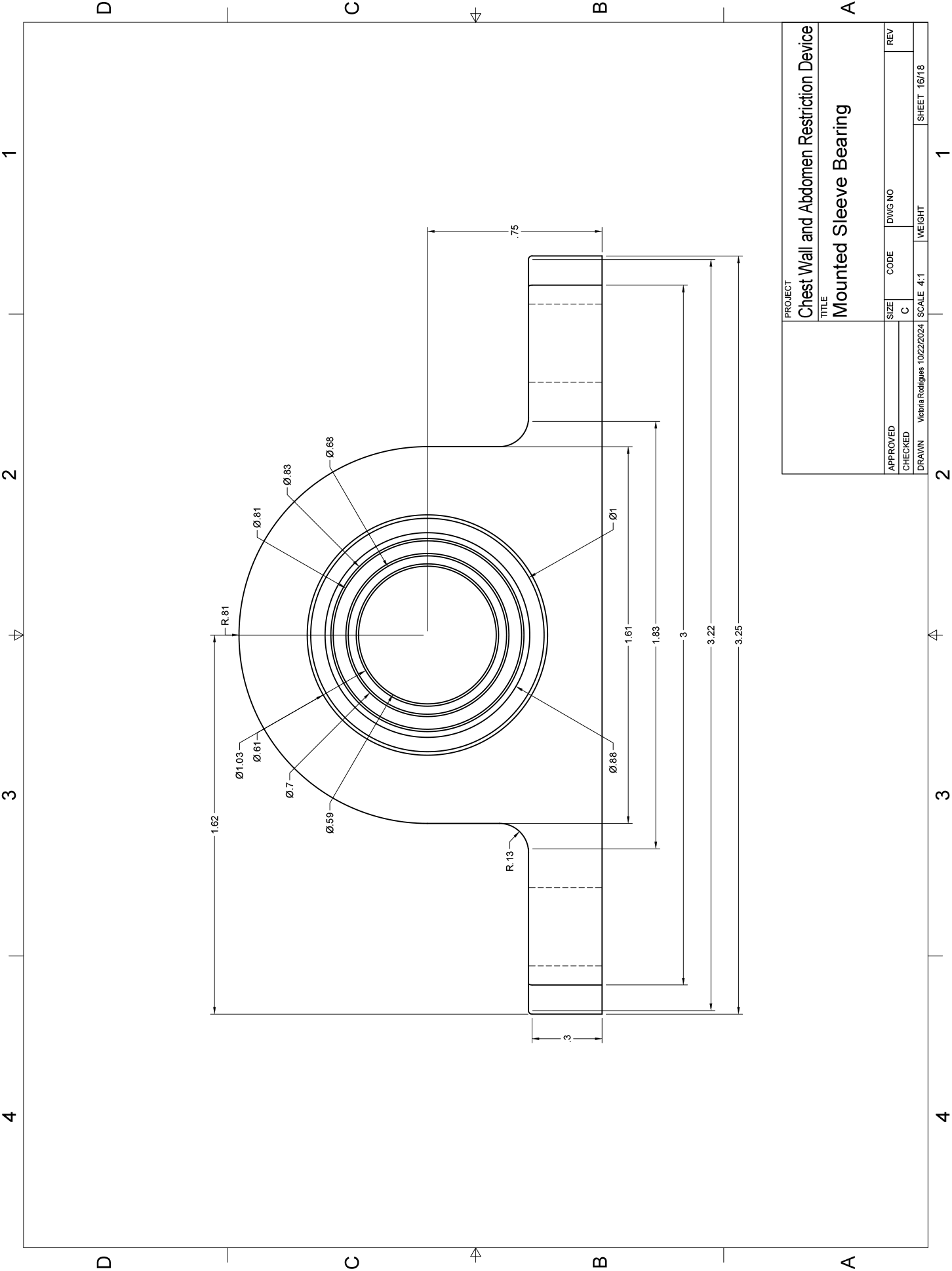

## Appendix Q Crank Handle

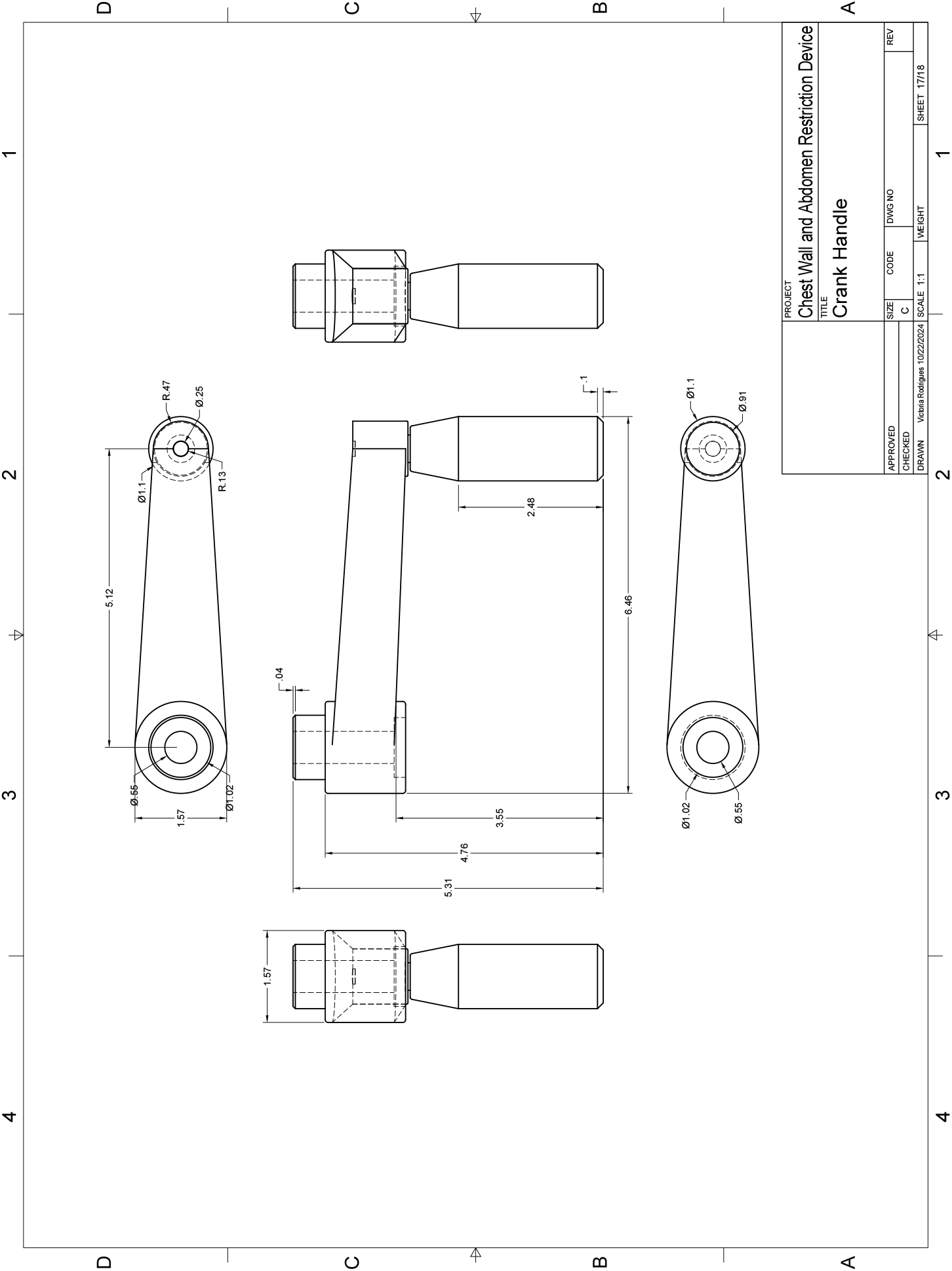

## Appendix R Presure Sensor

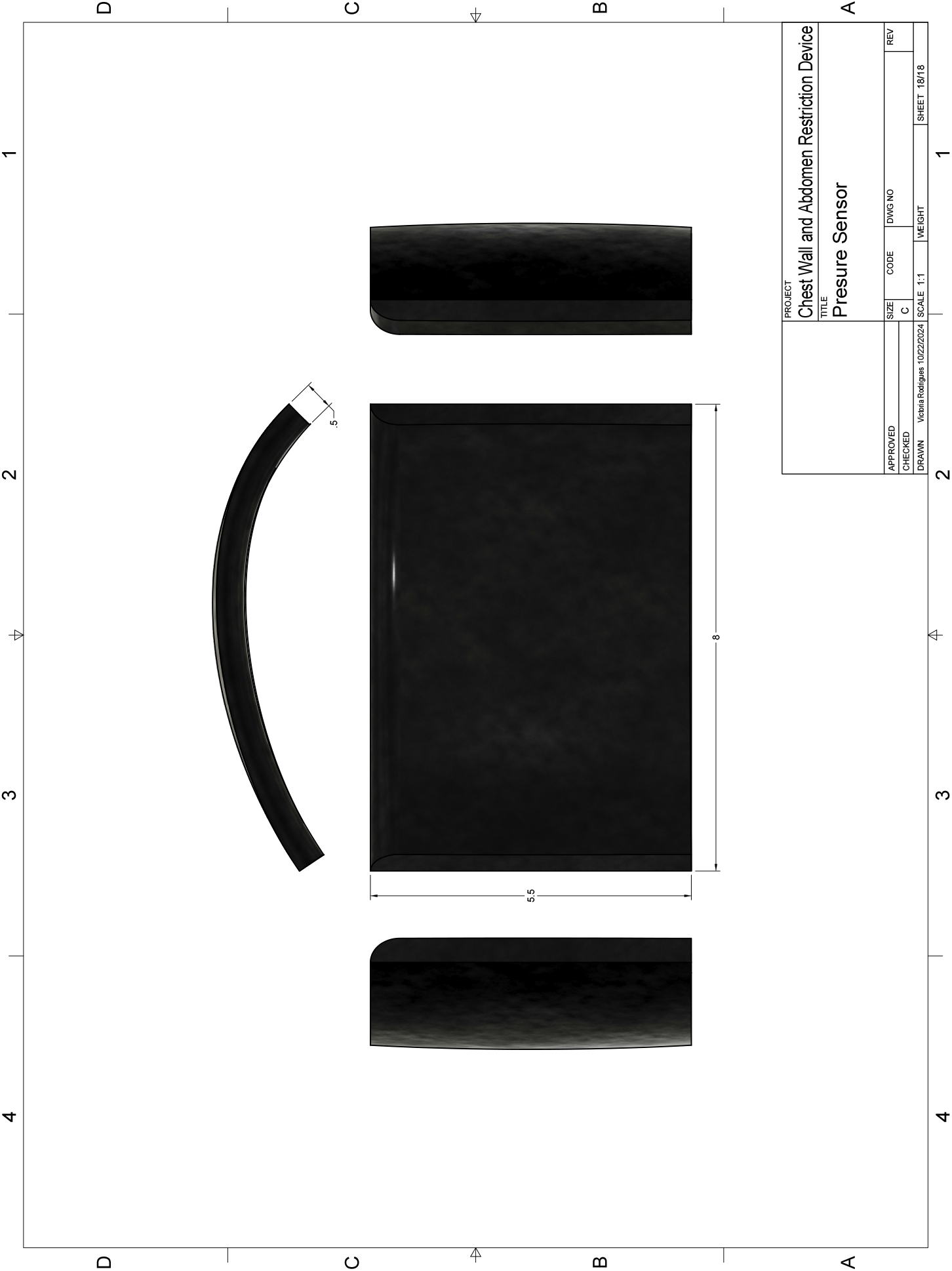

## Notes

This work was supported by the Office of Naval Research (grant number N00014-22-1-2653).

### Competing Interest Statement

The authors have declared no competing interest.

### Funding Statement

This work was supported by the Office of Naval Research (grant number N00014-22-1-2653).

### Author Declarations

The study procedures were approved by the Institutional Review Board of the University of Florida.

