## Appendix for "Chest Wall Restriction Device for Modeling Respiratory Challenges and Dysfunction"

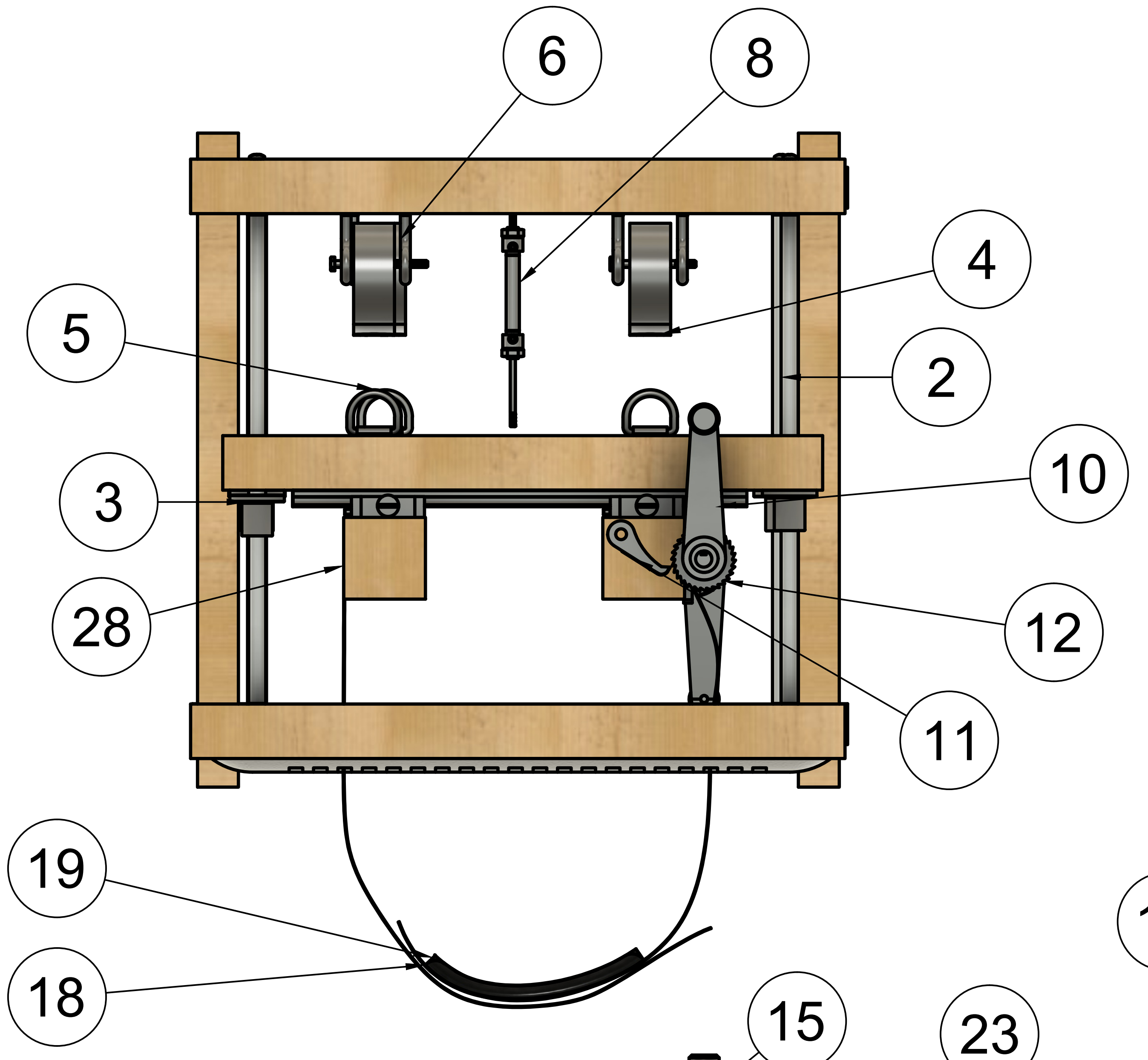


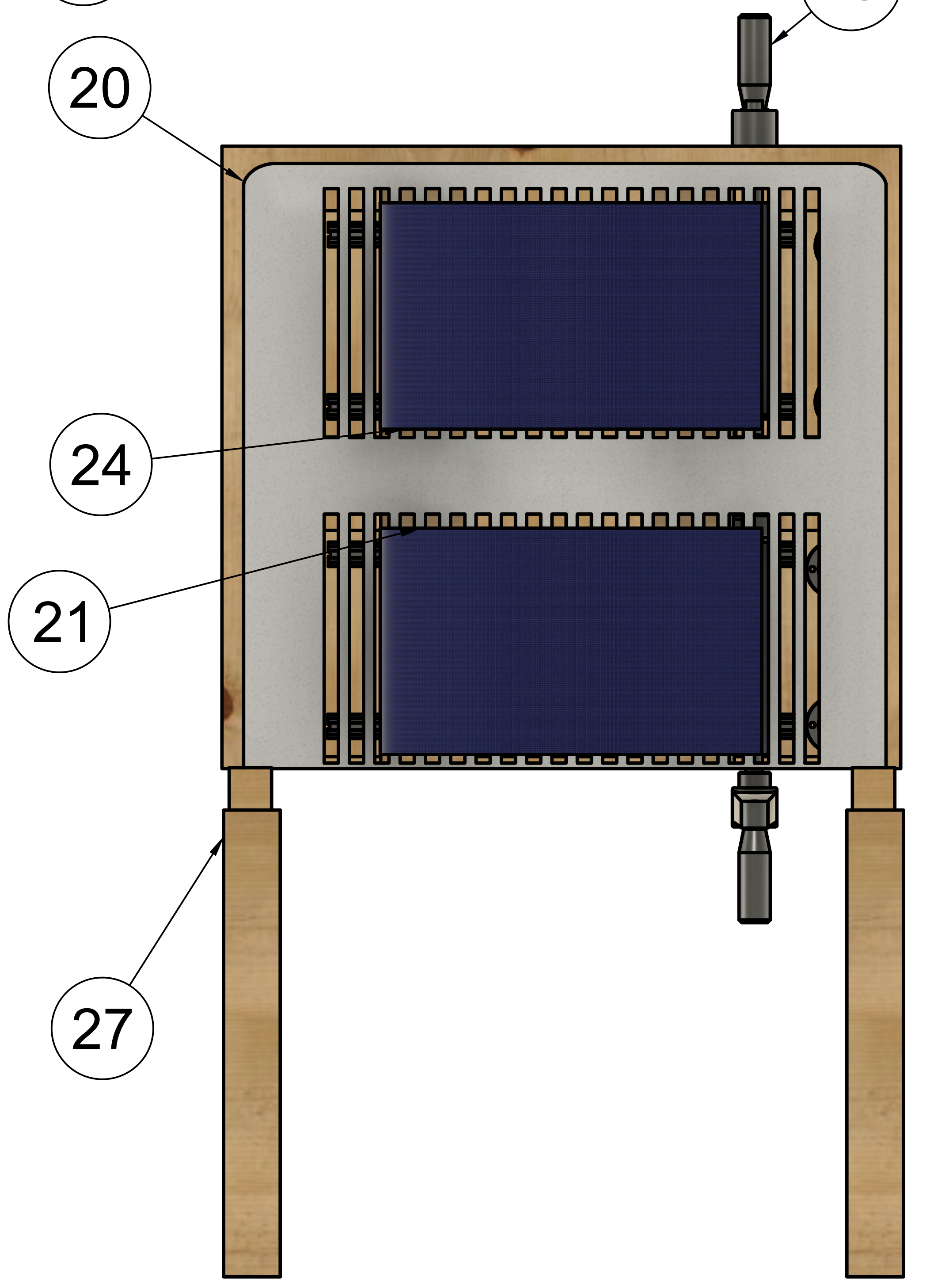


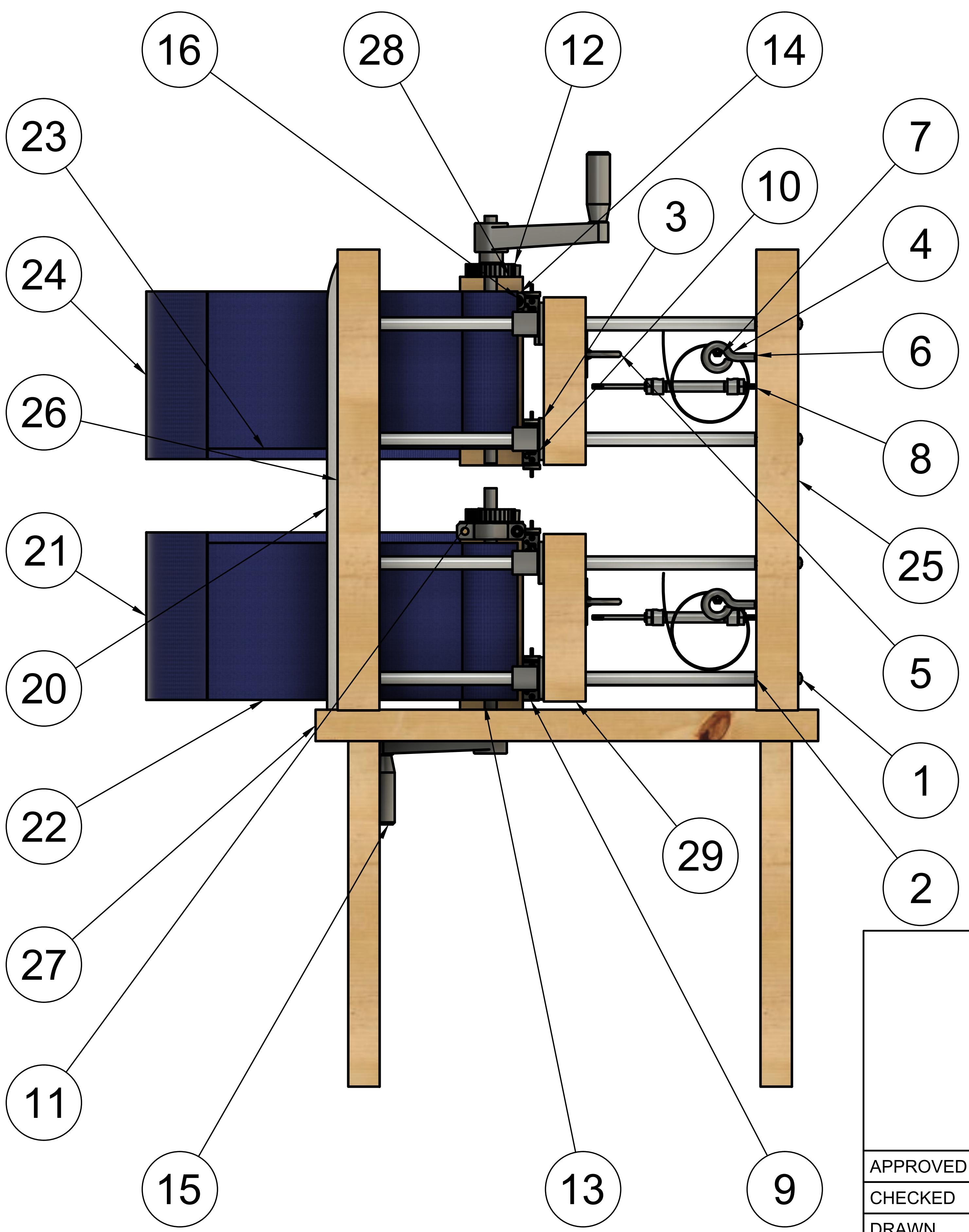


| PARTS LIST | | |
|---|---|---|
| ITEM | PART NUMBER | PART NAME |
| 1 | 91249A368 | PHILLIPS SCREWS |
| 2 | 6649K898 | TAPPED LINEAR MOTION SHAFT |
| 3 | 6483K54 | FLANGE-MOUNTED LINEAR BALL BEARING |
| 4 | 9293K905 | CONSTANT-FORCE SPRING |
| 5 | 3076T34 | TIE-DOWN RING |
| 6 | 9589T3 | ROUTING EYEBOLT |
| 7 | 92198A554 | HEX HEAD SCREW |
| 8 | LDI-619-100-A010S | LVDT |
| 9 | 3249K2 | LOCKING SLEEVE BEARING CARRIAGE |
| 10 | 9867K122 | GUIDE RAIL FOR BEARING CARRIAGE |
| 11 | 6283K83 | PAWL FOR METAL RATCHETING GEAR |
| 12 | 6283K78 | METAL RATCHETING GEAR |
| 13 | 1439K511_1045 | ROTARY SHAFT |
| 14 | 2820T56 | MOUNTED SLEEVE BEARING |
| 15 | 6546N17 | CRANK HANDLE |
| 16 | 91292A151 | SOCKET HEAD SCREW |
| 18 | PRESSURE SENSOR CHEST | PRESSURE SENSOR CHEST |
| 19 | PRESSURE SENSOR ABDOMEN | PRESSURE SENSOR ABDOMEN |
| 20 | FOAM PADDING | FOAM PADDING |
| 21 | RIGHT ABDOMINAL STRAP | RIGHT ABDOMINAL STRAP |
| 22 | LEFT ABDOMINAL STRAP | LEFT ABDOMINAL STRAP |
| 23 | LEFT CHEST STRAP | LEFT CHEST STRAP |
| 24 | RIGHT CHEST STRAP | RIGHT CHEST STRAP |
| 25 | | BACK BOARD |
| 26 | | FRONT BOARD |
| 27 | | BASE |
| 28 | | UPPER MIDDLE BOARD |
| 29 | | LOWER MIDDLE BOARD |

PROJECT
### Chest Wall and Abdomen Restriction Device

TITLE
#### Overview

| | | | | |
|---|---|---|---|---|
| APPROVED | SIZE C | CODE | DWG NO | REV |
| CHECKED | | | | |
| DRAWN Victoria Rodrigues 10/22/2024 | SCALE 1:5 | WEIGHT | SHEET 1/18 | |

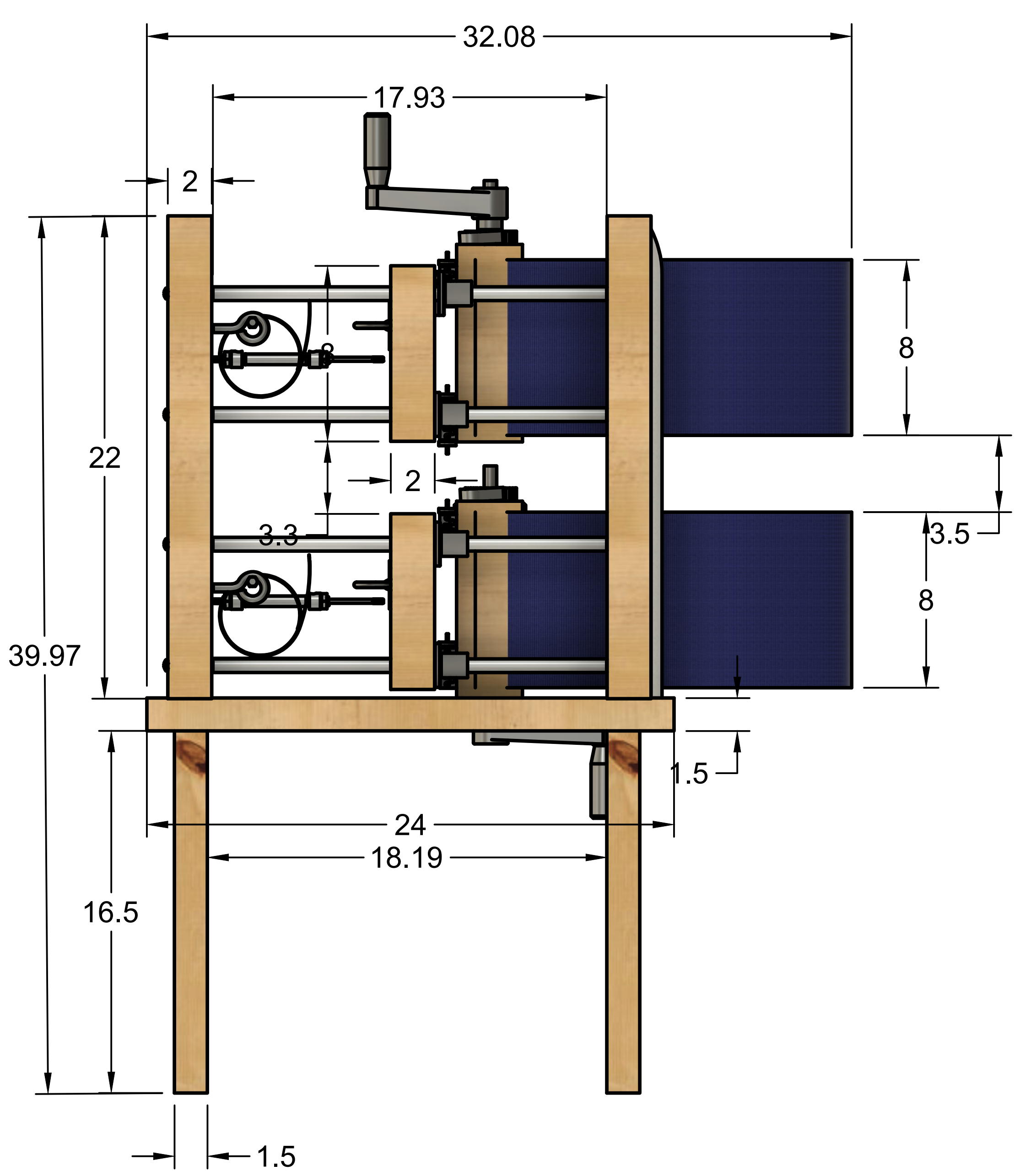


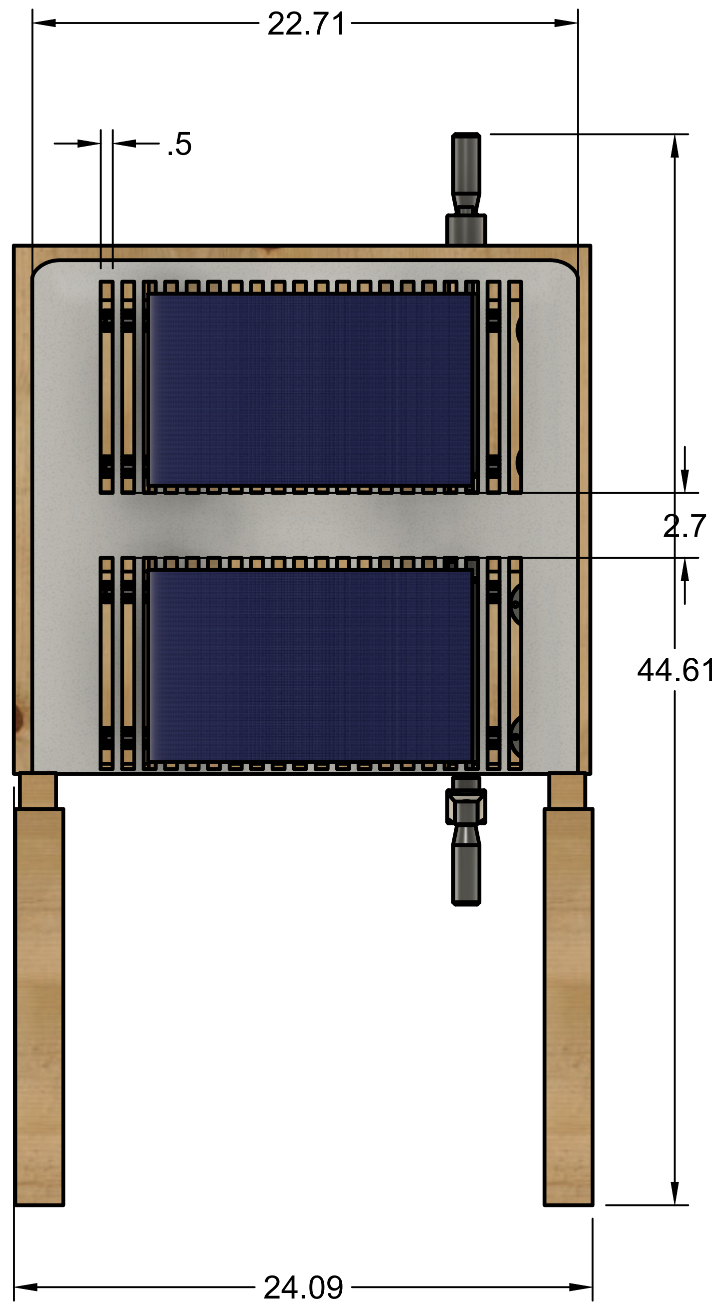


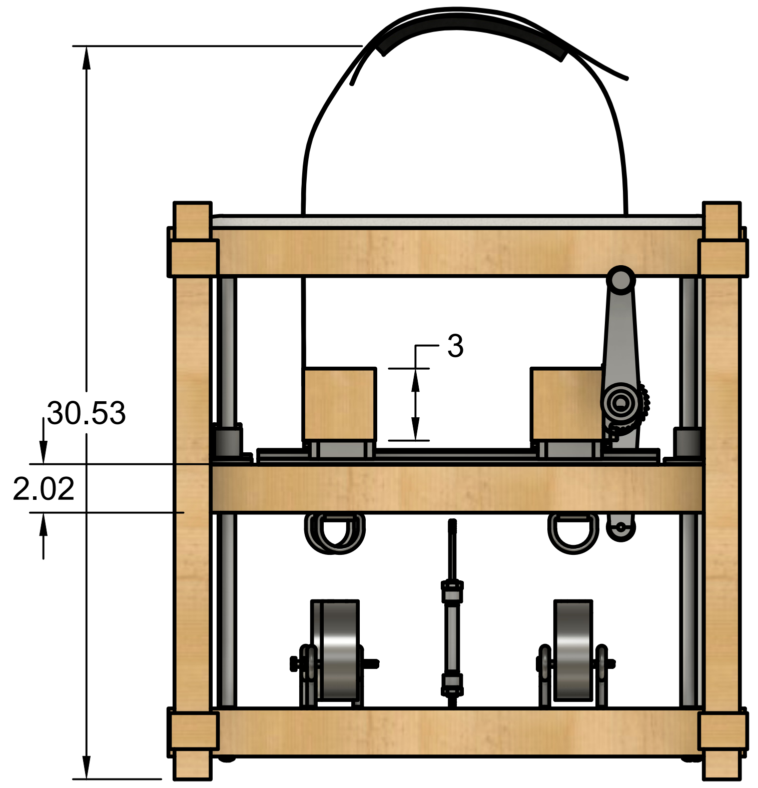


| | |
|---|---|
| PROJECT | Chest Wall and Abdomen Restriction Device |
| TITLE | Dimensions |

| | | | | |
|---|---|---|---|---|
| APPROVED | SIZE C | CODE | DWG NO | REV |
| CHECKED | | | | |
| DRAWN Victoria Rodrigues 10/22/2024 | SCALE 1:8 | WEIGHT | SHEET 2/18 | |

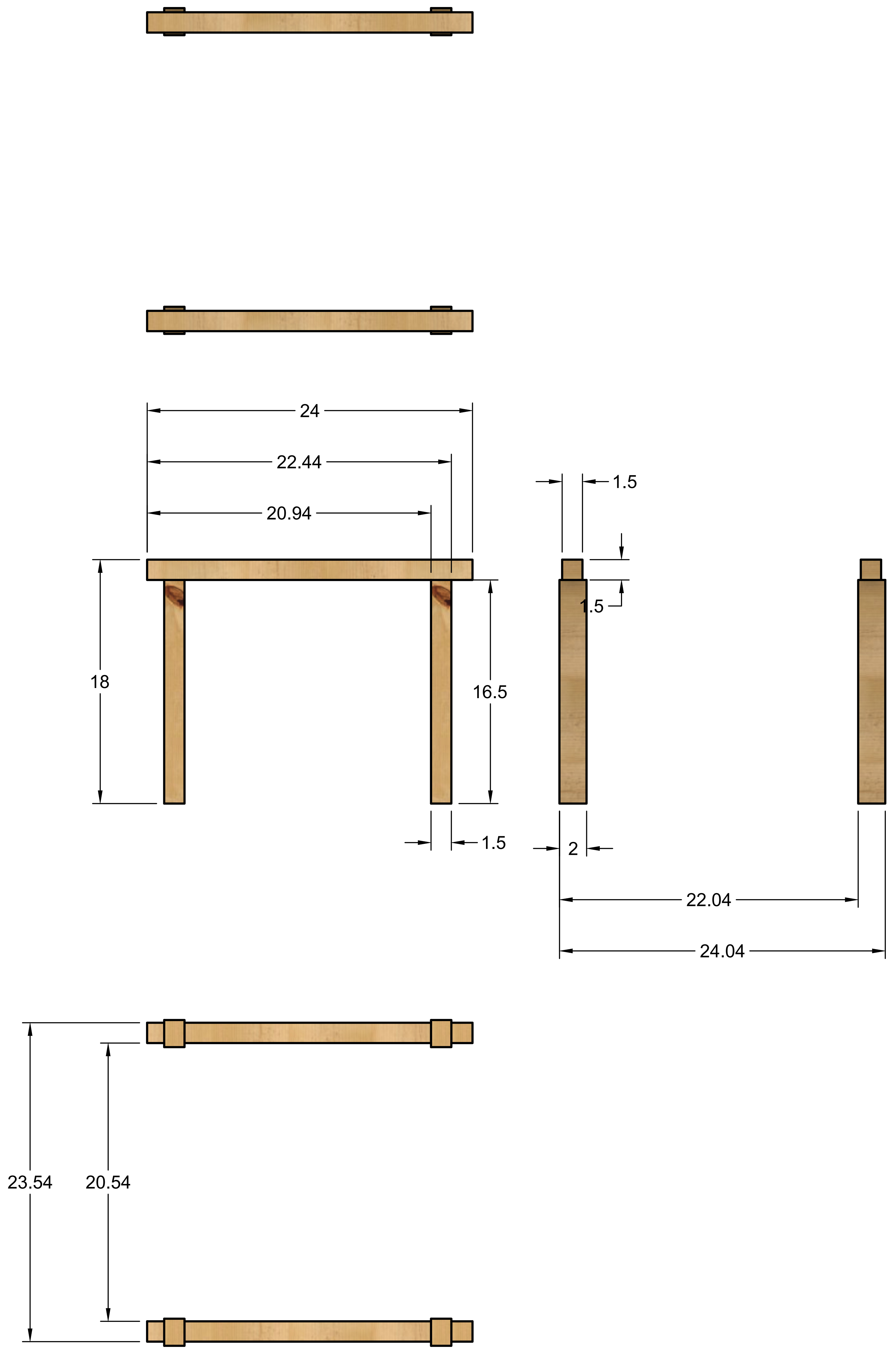


| | |
|---|---|
| PROJECT | Chest Wall and Abdomen Restriction Device |
| TITLE | Structural Base |
| APPROVED | |
| CHECKED | |
| DRAWN | Victoria Rodrigues 10/22/2024 |
| SIZE | C |
| CODE | |
| DWG NO | |
| REV | |
| SCALE | 1:8 |
| WEIGHT | |
| SHEET | 3/18 |

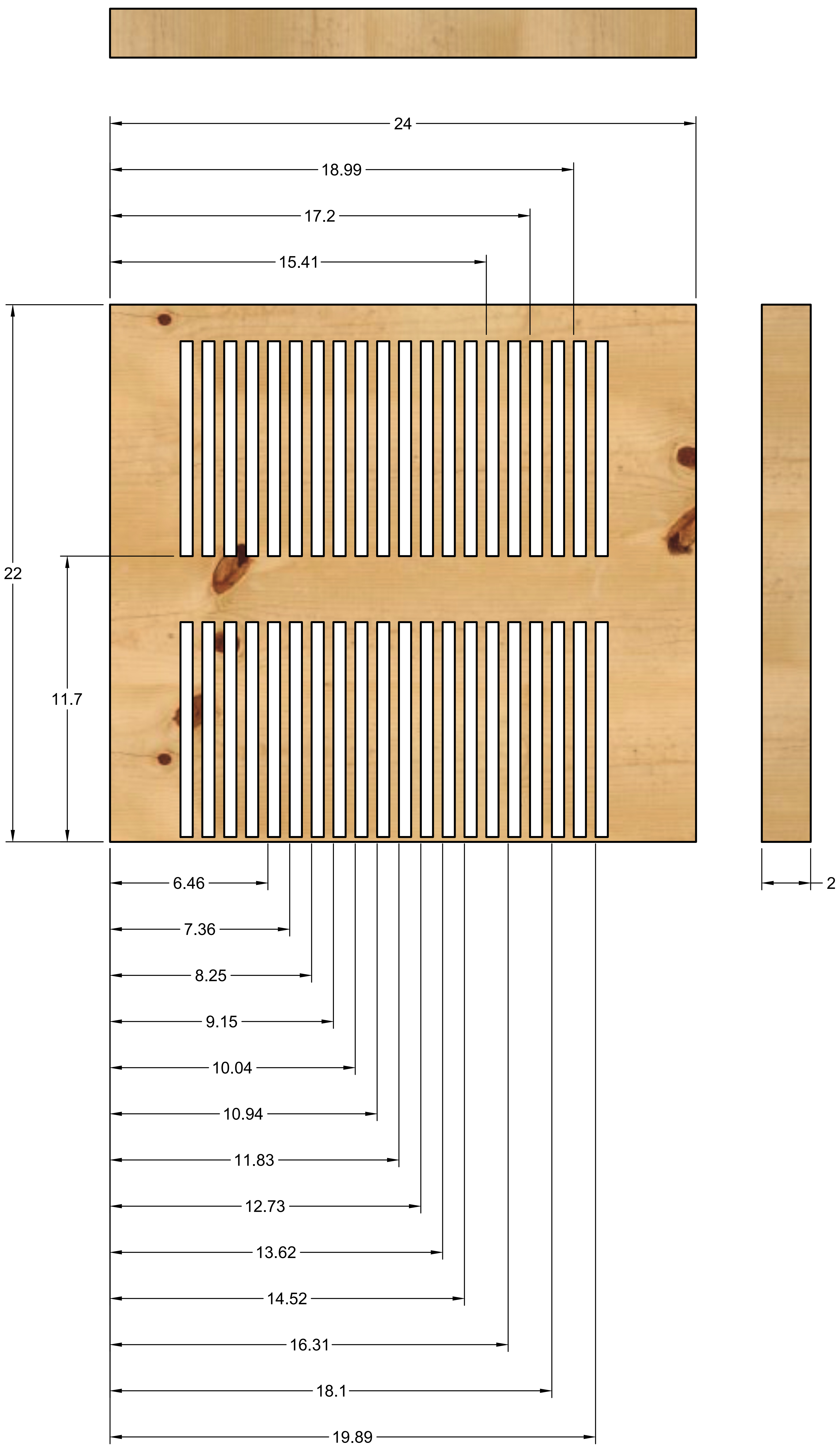


| | | | | |
|---|---|---|---|---|
| | PROJECT<br>Chest Wall and Abdomen Restriction Device | | | |
| | TITLE<br>Compression Surface | | | |
| APPROVED | SIZE | CODE | DWG NO | REV |
| CHECKED | C | | | |
| DRAWN Victoria Rodrigues 10/22/2024 | SCALE 1:4 | WEIGHT | SHEET 4/18 | |

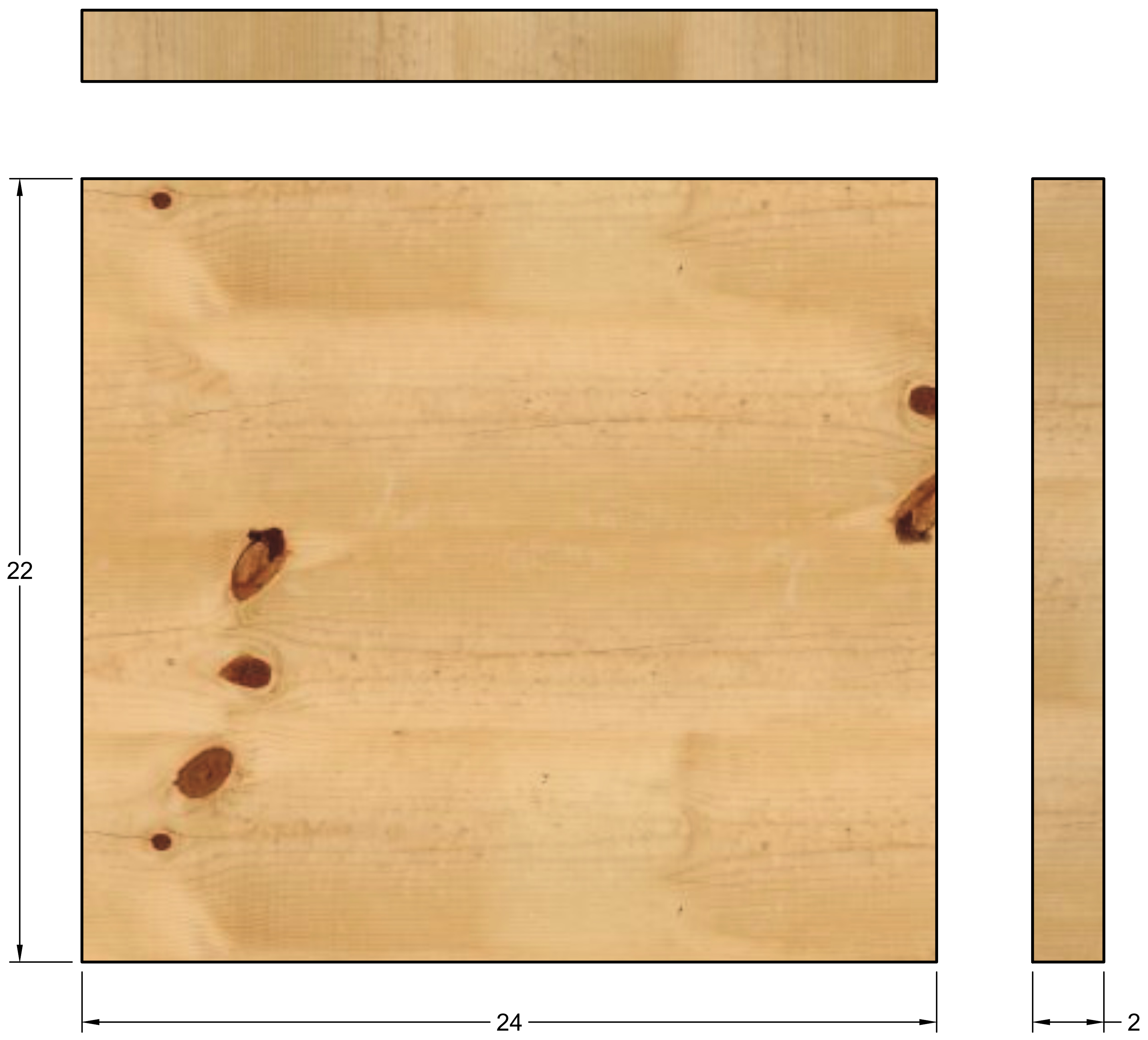


| | | | | | |
|---|---|---|---|---|---|
| | PROJECT<br>Chest Wall and Abdomen Restriction Device | | | | |
| | TITLE<br>Rear Support Board | | | | |
| APPROVED | SIZE | CODE | DWG NO | | REV |
| CHECKED | C | | | | |
| DRAWN Victoria Rodrigues 10/22/2024 | SCALE 1:4 | | WEIGHT | SHEET 5/18 | |

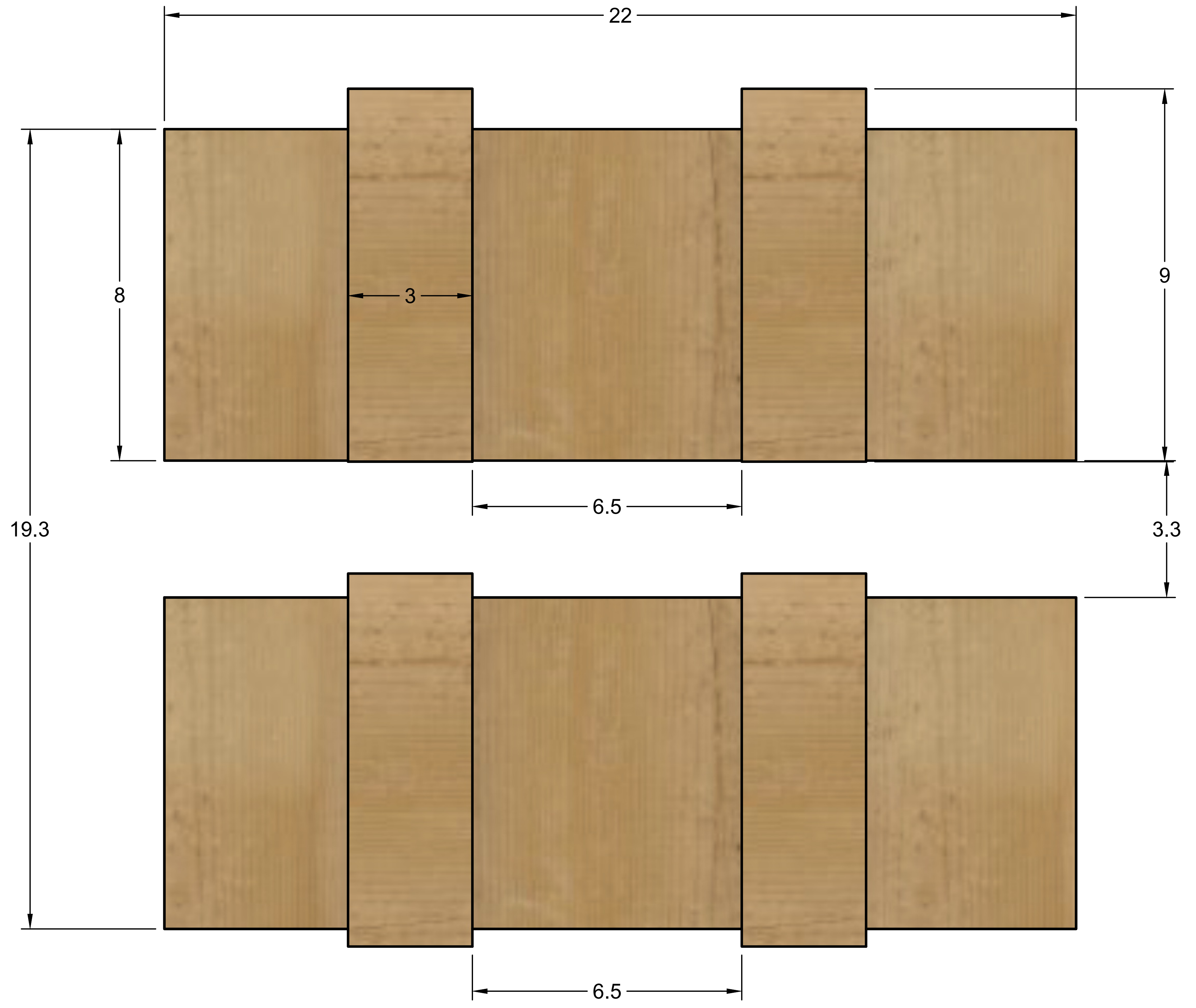


| | |
|---|---|
| | PROJECT<br>Chest Wall and Abdomen Restriction Device |
| | TITLE<br>Chest and Abdominal Restriction Boards |
| APPROVED | SIZE C · CODE · DWG NO · REV |
| CHECKED | |
| DRAWN Victoria Rodrigues 10/22/2024 | SCALE 1:6 · WEIGHT · SHEET 6/18 |

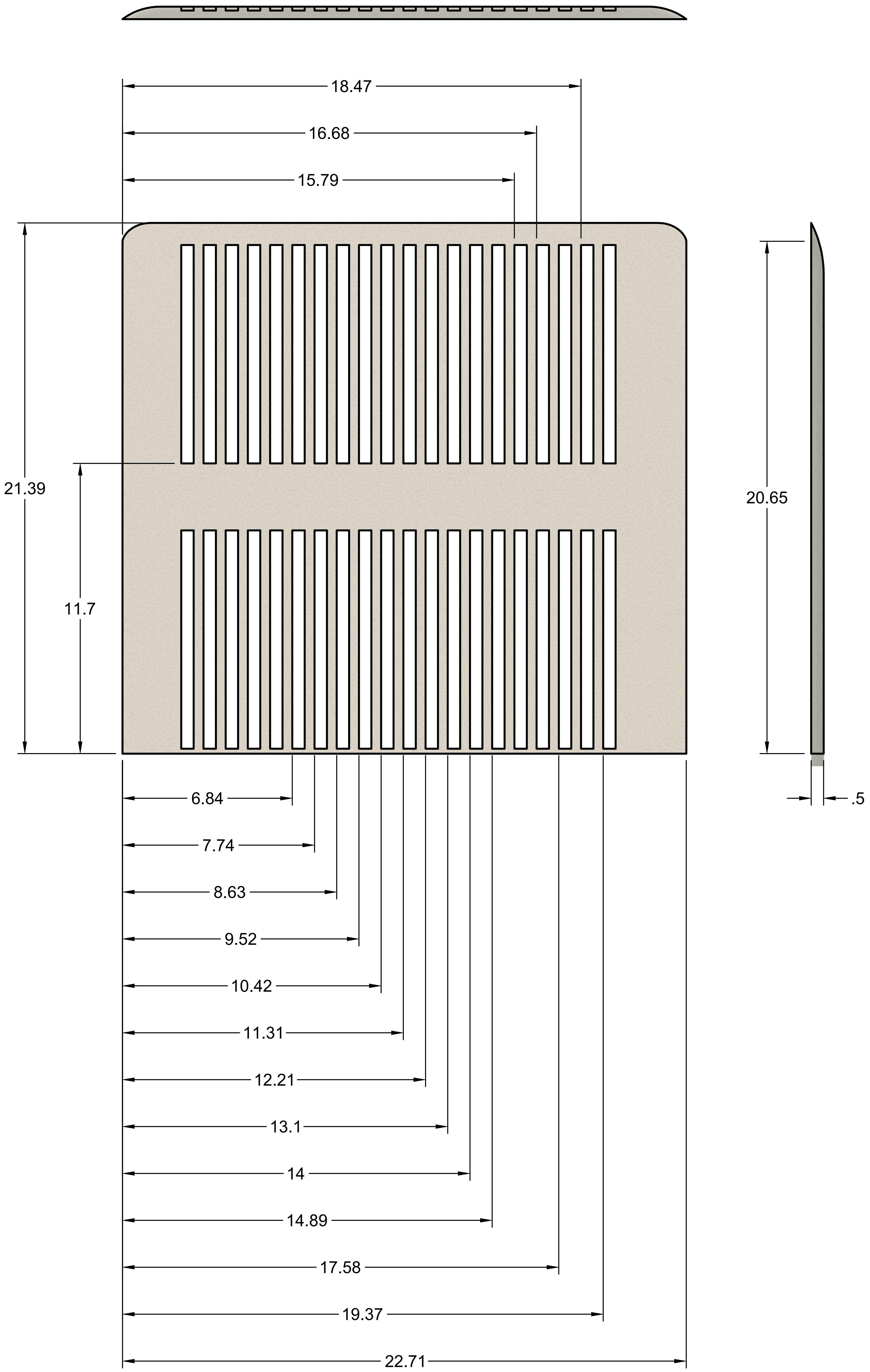


PROJECT
Chest Wall and Abdomen Restriction Device

TITLE
Foam Padding

| | | | | |
|---|---|---|---|---|
| APPROVED | SIZE C | CODE | DWG NO | REV |
| CHECKED | | | | |
| DRAWN Victoria Rodrigues 10/22/2024 | SCALE 1:4 | WEIGHT | SHEET 7/18 | |

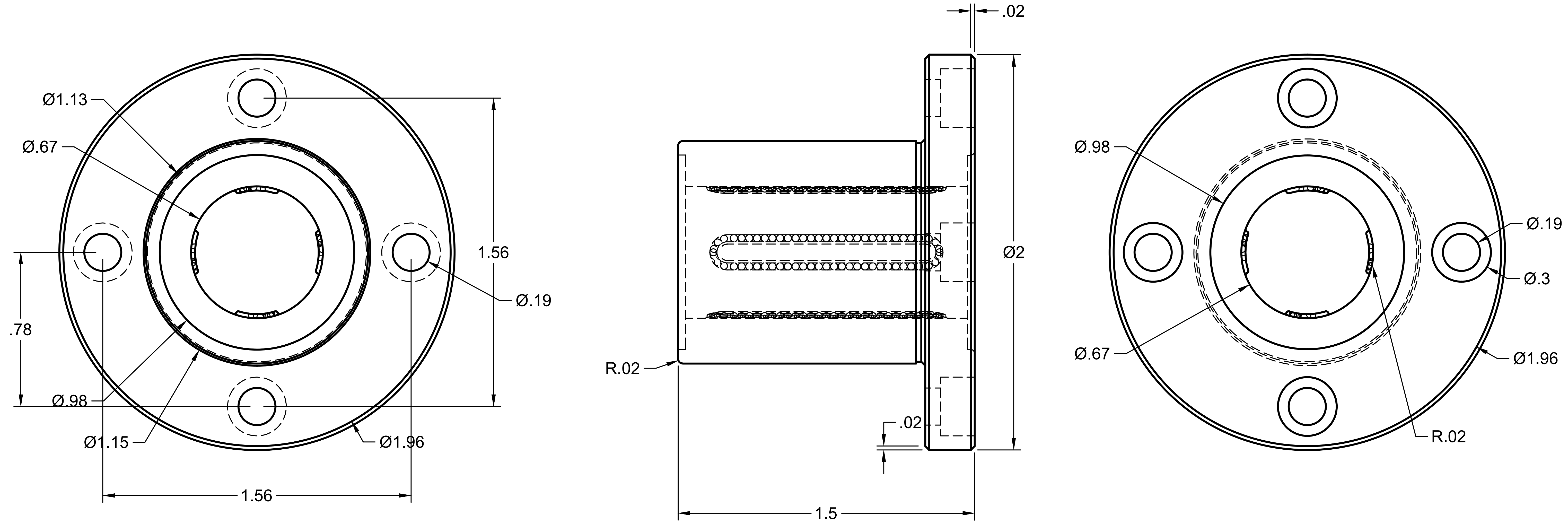


PROJECT
Chest Wall and Abdomen Restriction Device

TITLE
Flanged-Mounted Linear Ball Bearing

| | | | | |
|---|---|---|---|---|
| APPROVED | SIZE C | CODE | DWG NO | REV |
| CHECKED | | | | |
| DRAWN Victoria Rodrigues 10/22/2024 | SCALE 2:1 | WEIGHT | SHEET 8/18 | |

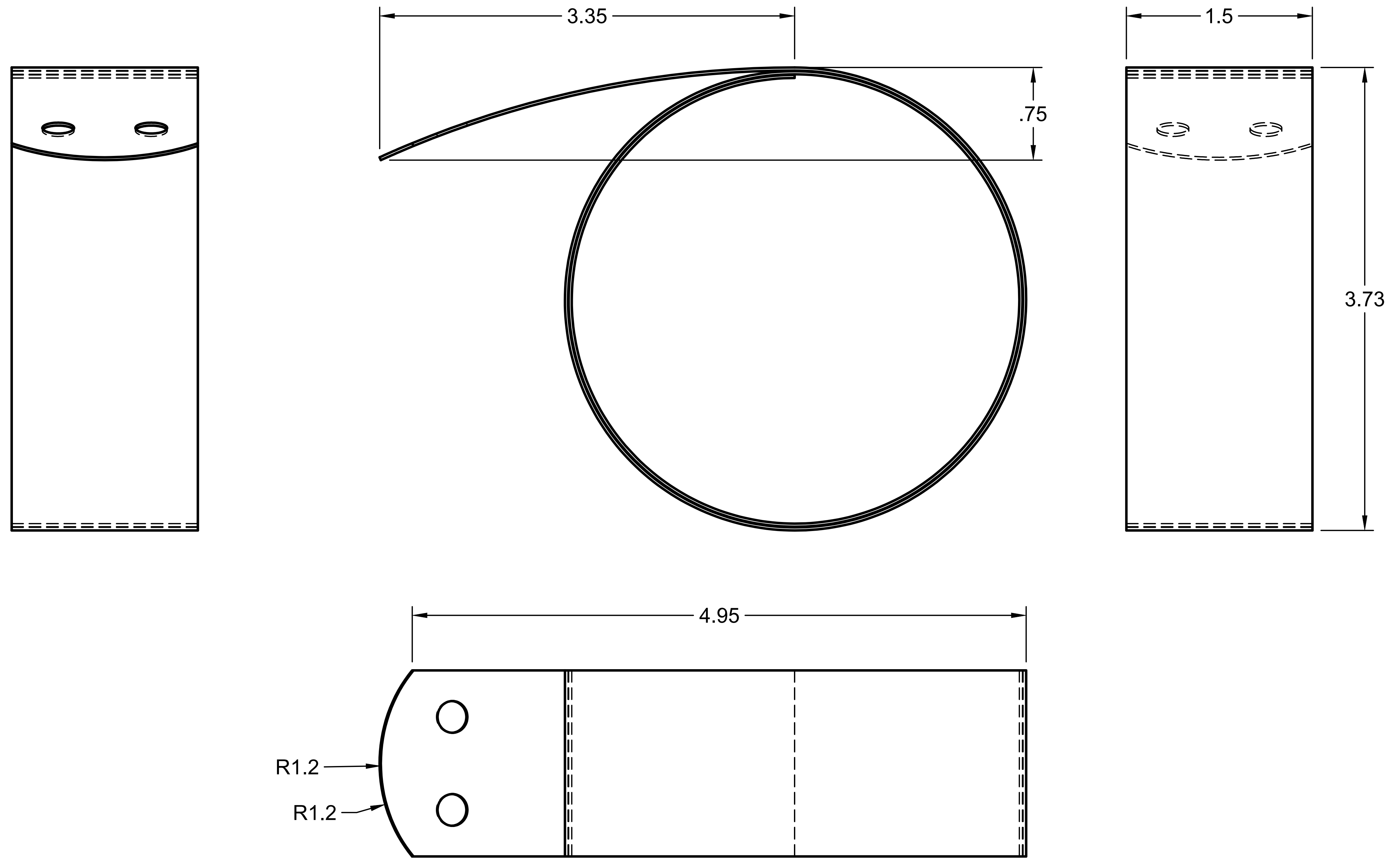


| | | | | |
|---|---|---|---|---|
| PROJECT | Chest Wall and Abdomen Restriction Device | | | |
| TITLE | Constant-Force Spring | | | |
| APPROVED | SIZE C | CODE | DWG NO | REV |
| CHECKED | | | | |
| DRAWN Victoria Rodrigues 10/22/2024 | SCALE 1:1 | WEIGHT | SHEET 9/18 | |

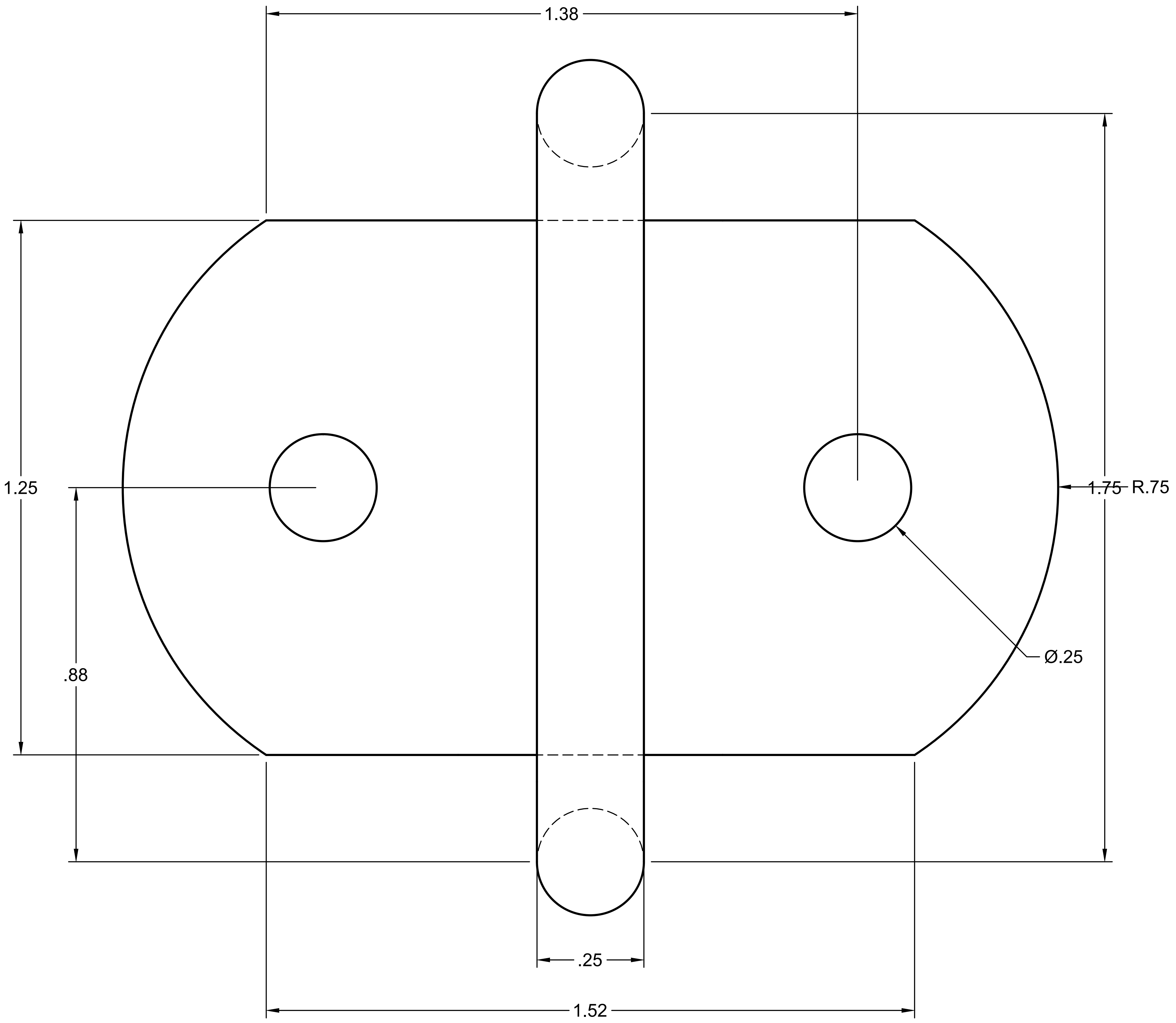


| | | | | |
|---|---|---|---|---|
| PROJECT<br>Chest Wall and Abdomen Restriction Device | | | | |
| TITLE<br>Tie-Down Ring | | | | |
| APPROVED | SIZE C | CODE | DWG NO | REV |
| CHECKED | | | | |
| DRAWN Victoria Rodrigues 10/22/2024 | SCALE 4:1 | WEIGHT | SHEET 10/18 | |

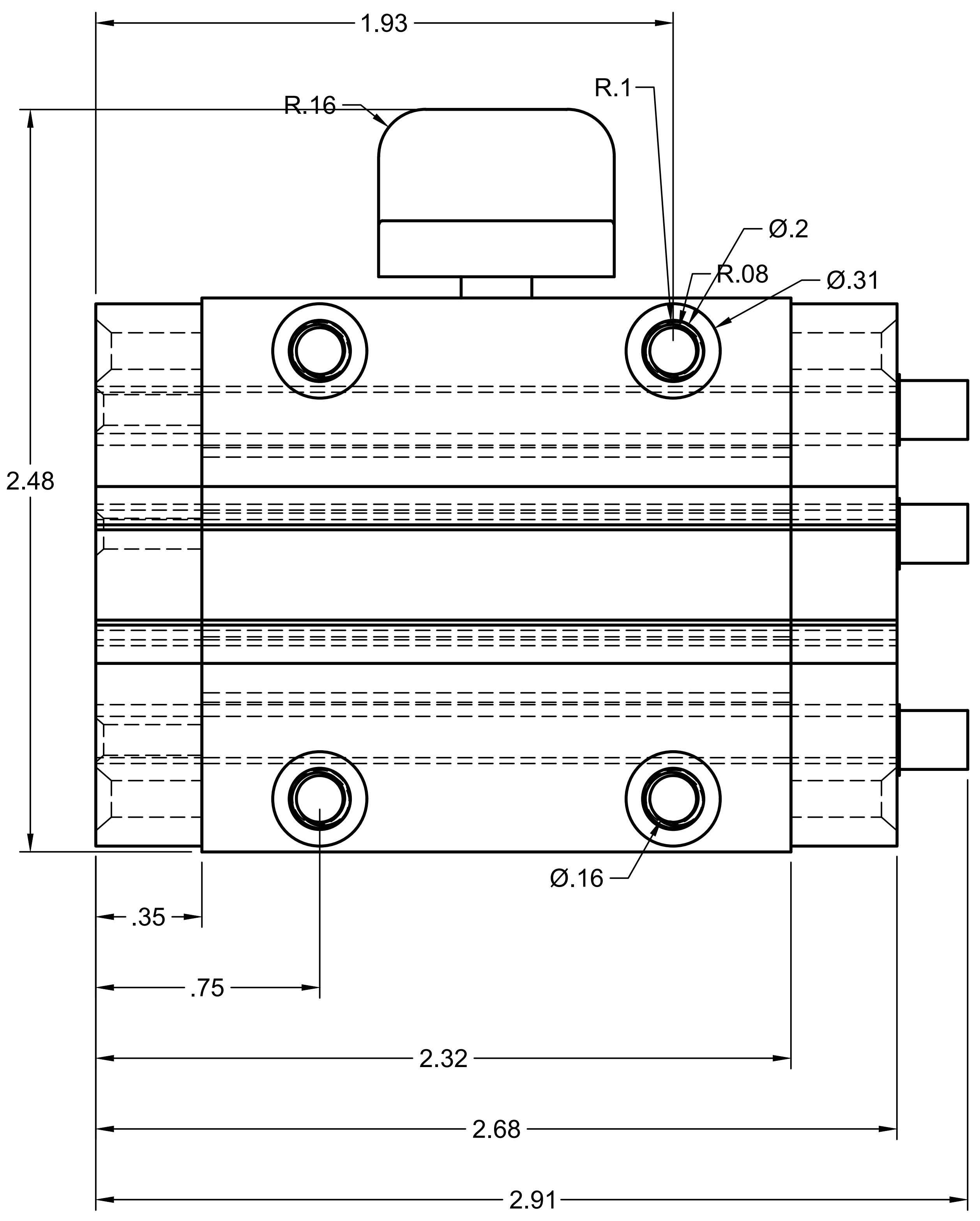


PROJECT

Chest Wall and Abdomen Restriction Device

TITLE

Locking Sleeve Bearing Carriage

| | | | | |
|---|---|---|---|---|
| APPROVED | SIZE | CODE | DWG NO | REV |
| CHECKED | C | | | |
| DRAWN Victoria Rodrigues 10/22/2024 | SCALE 2:1 | WEIGHT | | SHEET 11/18 |

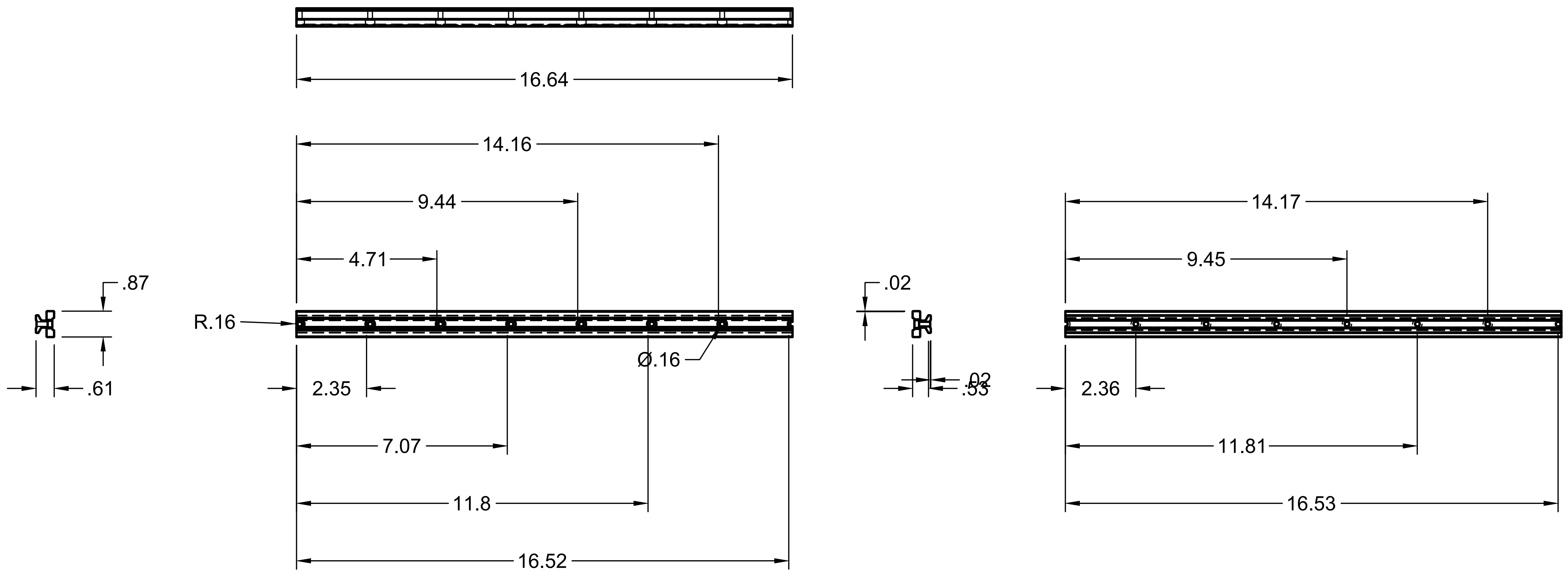


| | | | | | |
|---|---|---|---|---|---|
| | PROJECT Chest Wall and Abdomen Restriction Device | | | | |
| | TITLE Guide Rail for Bearing Carriage | | | | |
| APPROVED | SIZE C | CODE | DWG NO | | REV |
| CHECKED | | | | | |
| DRAWN Victoria Rodrigues 10/22/2024 | SCALE 1:4 | | WEIGHT | SHEET 12/18 | |

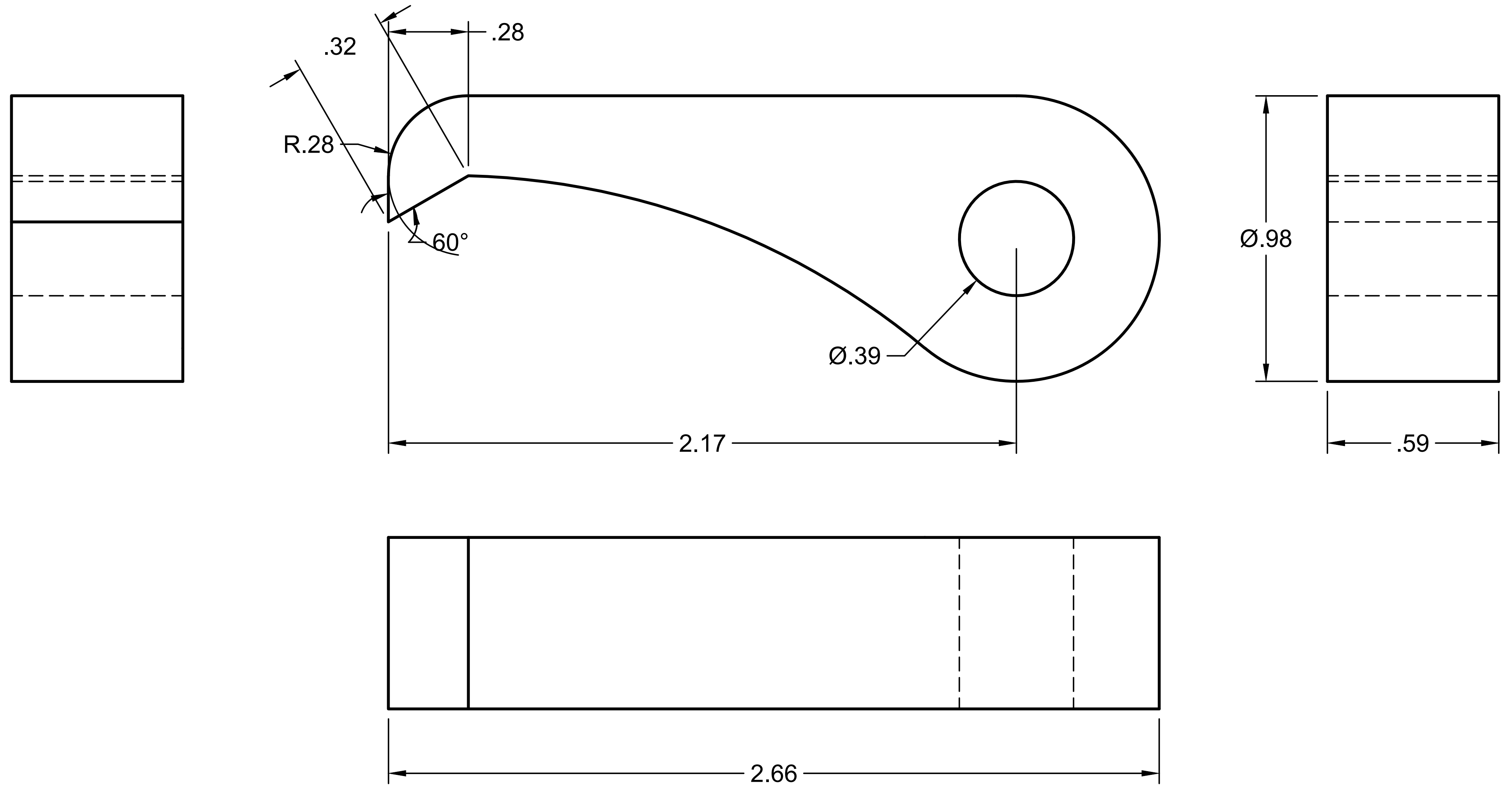


| | | | | |
|---|---|---|---|---|
| PROJECT Chest Wall and Abdomen Restriction Device | | | | |
| TITLE Pawl for Metal Ratcheting Gear | | | | |
| APPROVED | SIZE C | CODE | DWG NO | REV |
| CHECKED | | | | |
| DRAWN Victoria Rodrigues 10/22/2024 | SCALE 2:1 | WEIGHT | SHEET 13/18 | |

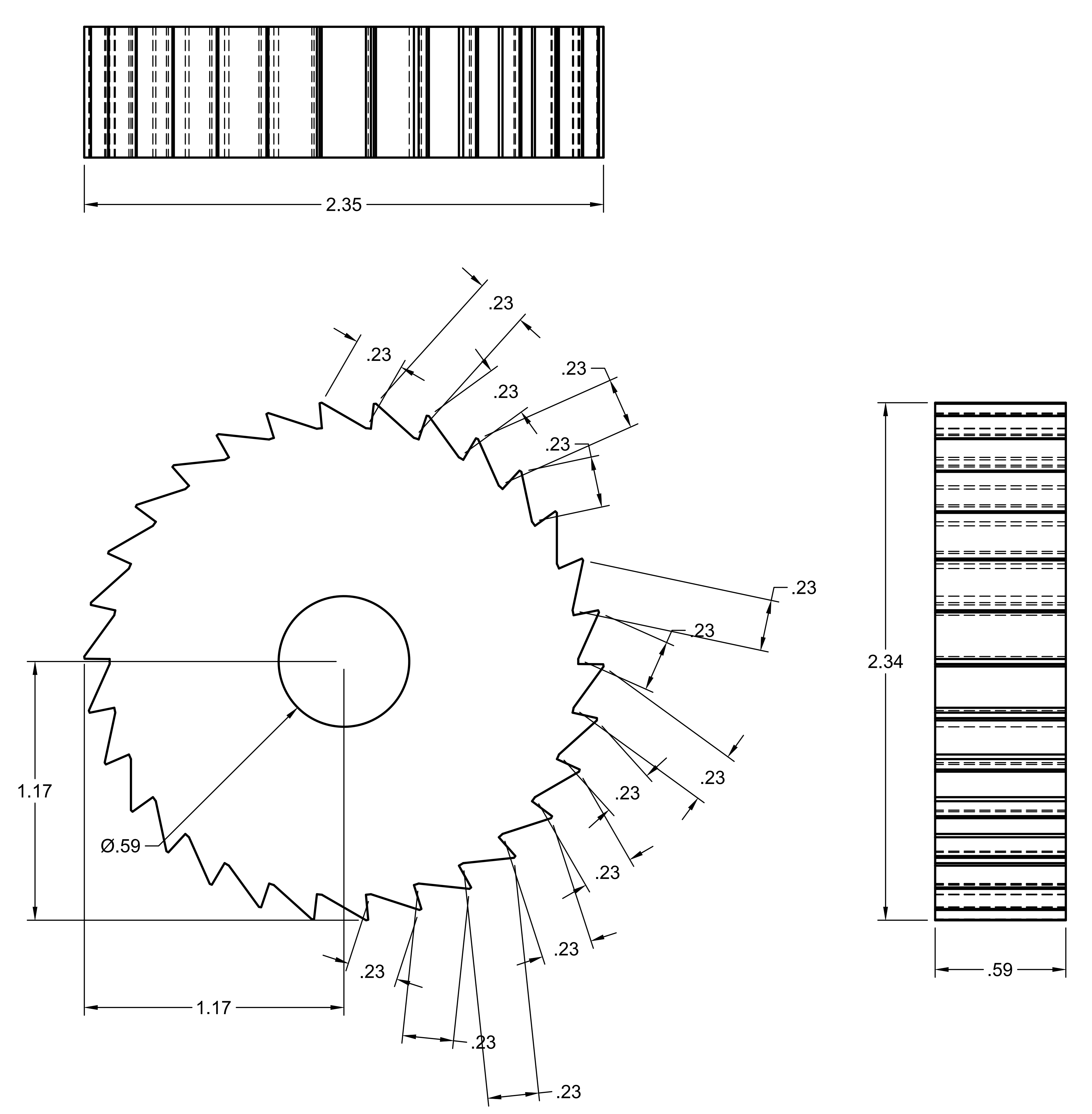


| | | | | |
|---|---|---|---|---|
| | PROJECT<br>Chest Wall and Abdomen Restriction Device | | | |
| | TITLE<br>Metal Ratcheting Gear | | | |
| APPROVED | SIZE<br>C | CODE | DWG NO | REV |
| CHECKED | | | | |
| DRAWN Victoria Rodrigues 10/22/2024 | SCALE 2:1 | WEIGHT | | SHEET 14/18 |

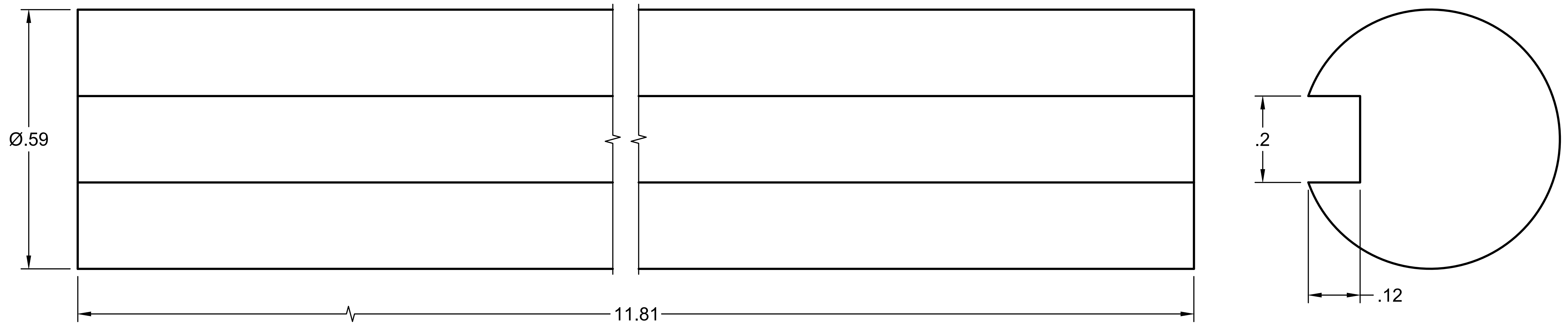


| | | | | | |
|---|---|---|---|---|---|
| | PROJECT Chest Wall and Abdomen Restriction Device | | | | |
| | TITLE Rotary Shaft | | | | |
| APPROVED | SIZE C | CODE | DWG NO | | REV |
| CHECKED | | | | | |
| DRAWN Victoria Rodrigues 10/22/2024 | SCALE 4:1 | | WEIGHT | SHEET 15/18 | |

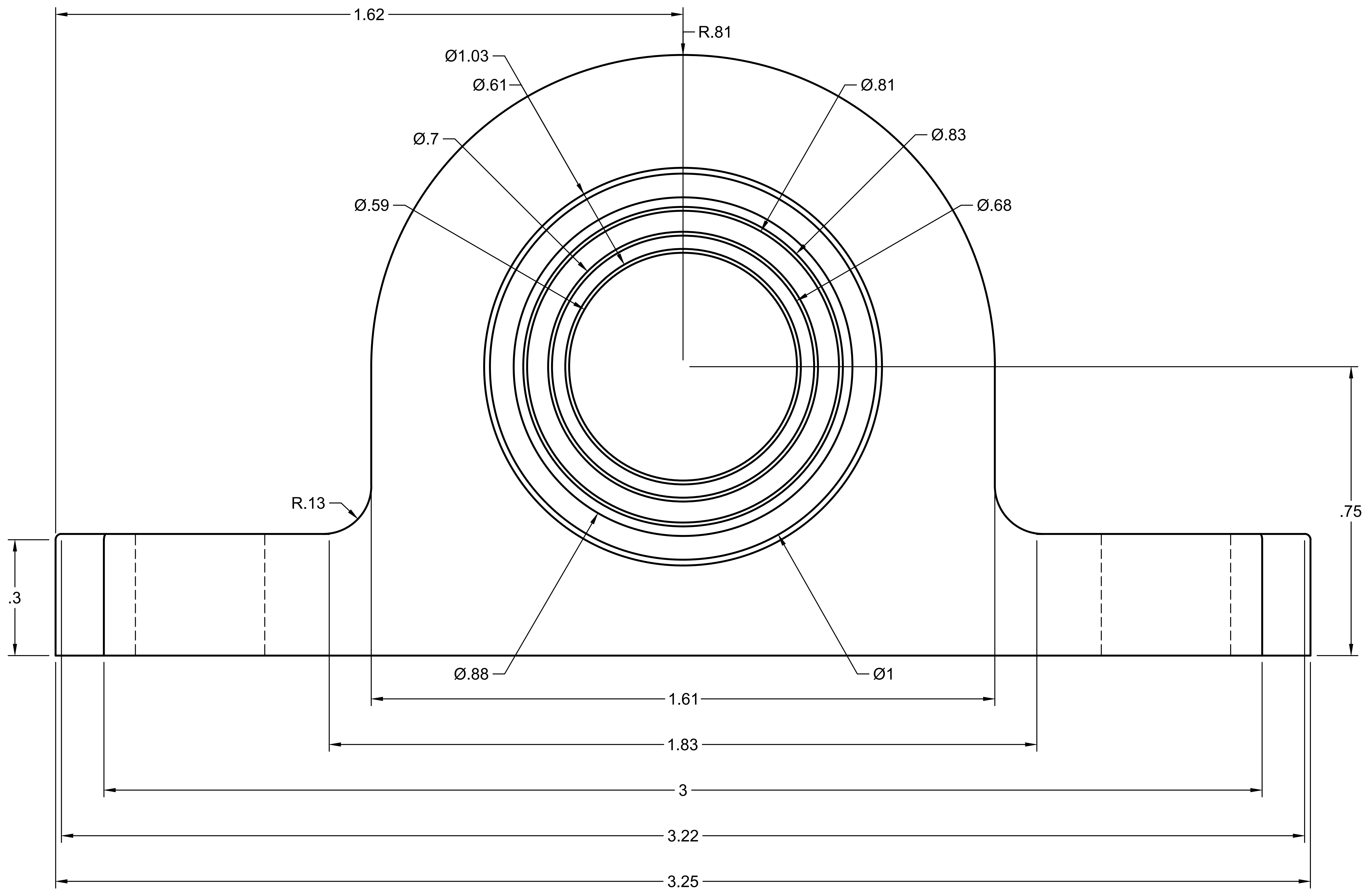


| | |
|---|---|
| PROJECT | Chest Wall and Abdomen Restriction Device |
| TITLE | Mounted Sleeve Bearing |
| APPROVED | |
| CHECKED | |
| DRAWN | Victoria Rodrigues 10/22/2024 |
| SIZE | C |
| CODE | |
| DWG NO | |
| REV | |
| SCALE | 4:1 |
| WEIGHT | |
| SHEET | 16/18 |

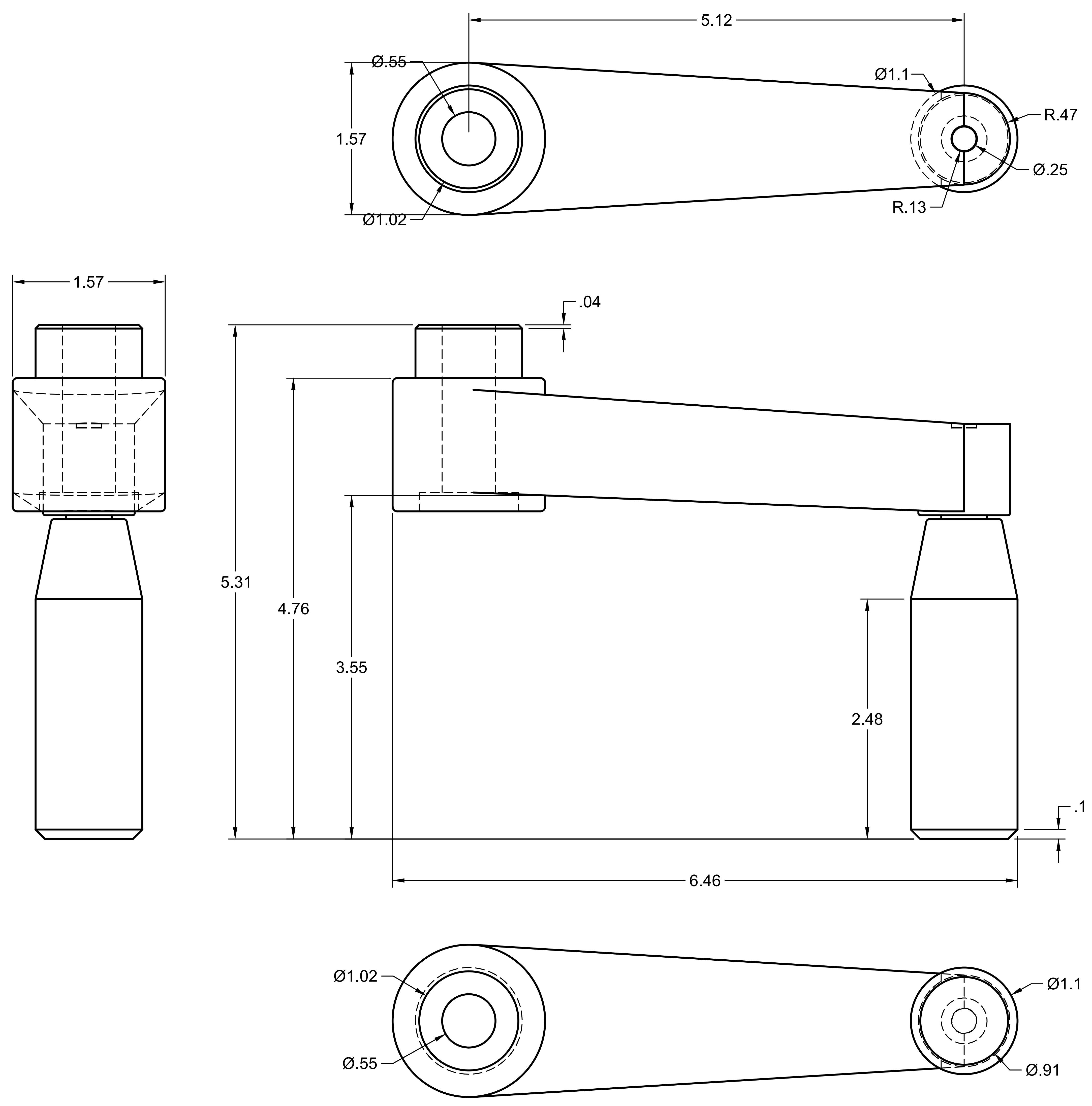


PROJECT
Chest Wall and Abdomen Restriction Device
TITLE
Crank Handle

| | | | | |
|---|---|---|---|---|
| APPROVED | SIZE C | CODE | DWG NO | REV |
| CHECKED | | | | |
| DRAWN Victoria Rodrigues 10/22/2024 | SCALE 1:1 | WEIGHT | SHEET 17/18 | |

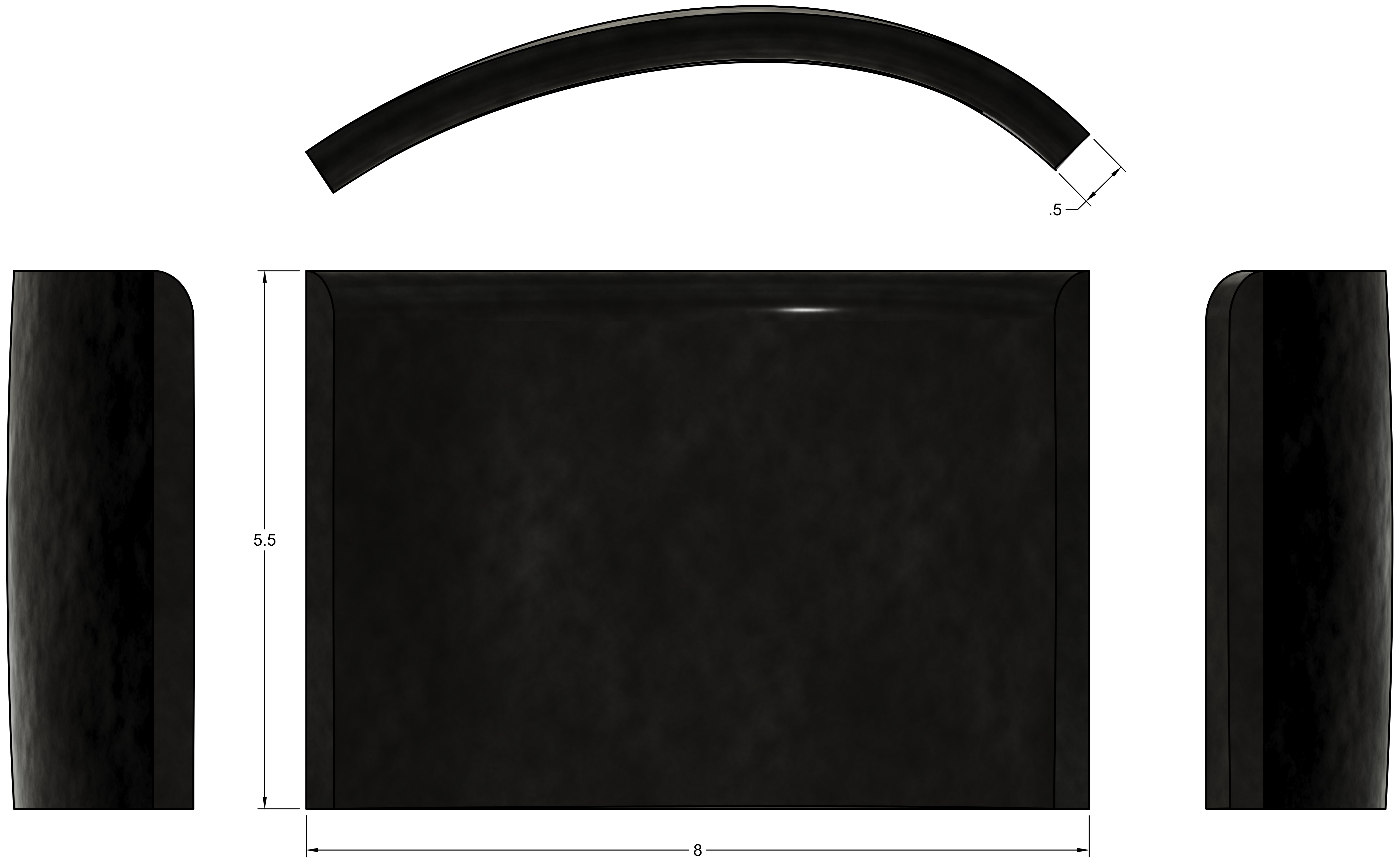


| | |
|---|---|
| PROJECT | Chest Wall and Abdomen Restriction Device |
| TITLE | Presure Sensor |

| | | | | |
|---|---|---|---|---|
| APPROVED | SIZE C | CODE | DWG NO | REV |
| CHECKED | | | | |
| DRAWN Victoria Rodrigues 10/22/2024 | SCALE 1:1 | WEIGHT | SHEET 18/18 | |
